# Evaluation of different stool extraction methods for metabolomics measurements in human fecal samples

**DOI:** 10.1101/2020.10.12.20209767

**Authors:** Vanessa Erben, Gernot Poschet, Petra Schrotz-King, Hermann Brenner

## Abstract

**Background:** Measurement of metabolomics in human stool samples is of great interest for a broad range of applications in biomedical research including early detection of colorectal neoplasms. However, due to the complexity of metabolites there is no consensus on how to process samples for stool metabolomics measurements to obtain a broad coverage of hydrophilic and hydrophobic substances.

**Methods:** We used frozen stool samples (50mg) from healthy study participants. Stool samples were processed after thawing using 8 different processing protocols and different solvents. Metabolites were measured afterwards using the MxP® Quant 500 kit (Biocrates). The best performing protocol was subsequently applied to compare stool samples of participants with different dietary habits.

**Results:** In this study, we were able to determine up to 340 metabolites of various chemical classes extracted from stool samples of healthy study participants with 8 different protocols. Polar metabolites such as amino acids could be measured with each method while other metabolite classes, particular lipid species, are more dependent on the solvent or combination of solvents used. Only a small number of triglycerides or acylcarnitines were detected in human feces. Extraction efficiency was higher for protocols using isopropanol or those using ethanol or methanol and MTBE including an evaporation and concentration step than for other protocols. We detected significant fecal metabolite differences between vegetarians, semi-vegetarians and non-vegetarians.

**Conclusion:** For the evaluation of metabolites in fecal samples we found protocols using solvents like isopropanol and those using ethanol or methanol and MTBE including an evaporation and concentration step to be superior over others tested in this study.

## Introduction

Measurement of metabolomics in human stool samples is of great interest for a broad range of applications in biomedical research including early detection of colorectal cancer (CRC) as a non-invasive alternative to colonoscopy. Metabolomics might be promising for this purpose as metabolites are closely related to the phenotype and depict current metabolic processes happening in an organism. Stool is directly associated with the gut and may reflect changes in metabolism very early through its transit in the gut (1).

Fecal mass consists to a great proportion of water and bacteria but also food components or metabolites (2). Fecal samples show great variability in their material content and characteristics, which makes it difficult to standardize the process from collection to processing and analysis including the analytical platform (3). The latter defines also the sensitivity of the analysis and the type of analytes available for analyses. Apart from the afore mentioned differences of stool samples, metabolic changes in stool might be derived directly from the development of cancer or precancerous cells or from a change in the gut microbiota which both result in a distinct metabolic phenotype that might be characteristic for the disease (1). The metabolic profile of CRC or its precursors may help in the understanding of disease development, progression and early detection (4).

Some studies have already found fecal metabolomics biomarkers for early detection of CRC but metabolite selection strongly varied (1, 5-7) and as different studies were using different processing methods, no direct comparison is possible. There is no consensus how to process stool samples for metabolomics measurements to get reliable and reproducible results. A review by Deda et al. focused on the existing various stool preparation protocols and found the metabolites to be dependent on the extraction method (8). In this study we used the MxP® Quant 500 Kit (Biocrates Life Sciences AG, Innsbruck, Austria) to determine and quantify a very broad range of metabolite classes in human feces. In total, we used 8 different stool preparation protocols to assess the best coverage for stool metabolite profiles. The protocol yielding the highest multitude of extracted metabolites was used to analyze and compare additional stool samples of healthy study participants with different dietary habits.

## Subjects and Methods

### Study design

The GEKKO study (Gebt dem Krebs keine Chance – Onkocheck) is an ongoing study in southwest Germany including people participating in screening colonoscopy (Arm A) or with diagnosed primary cancer (Arm B). The study was approved by the ethics committees of the Medical Faculty Heidelberg and of the physicians’ boards of Baden-Württemberg and Rhineland Palatinate. The GEKKO study Arm A was designed to evaluate novel early detection markers of CRC. People undergoing a screening colonoscopy at a gastroenterology practice in or around Heidelberg, Germany, who are over the age of 30 years, with no history of CRC, no inflammatory bowel disease, no colonoscopy within the last 5 years and speaking and understanding the German language are invited to participate.

After written informed consent was received, participants are asked to fill in a questionnaire regarding lifestyle and demographic data and to provide blood, stool, saliva and urine samples for research purposes prior to colonoscopy. Biosamples are processed and then stored at -80°C until needed. Colonoscopy reports are provided from the physicians to the study center. Participants are grouped according to their most advanced finding at colonoscopy. From the individuals with no polyps or any findings at colonoscopy, those with incomplete colonoscopy (Coecum not reached) or poor bowel preparation were excluded. For this analysis, 3 participants of the GEKKO study Arm A were selected between the age of 50 and 79 years with no polyps or any findings at colonoscopy to test 8 different stool preparation protocols and to define the best analytical outcome.

In a further step, stool samples from additional healthy GEKKO participants (n = 18) of approximately the same age (50 – 65 years) with different dietary habits (vegetarians, semi-vegetarians, and non-vegetarians) were processed with the protocol that performed best with respect to numbers of detected metabolites and sample handling, and results were compared between groups. Information on dietary habits of the study participants was extracted from the questionnaire. Vegetarians were defined as never eating meat, processed meat and poultry. Semi-vegetarians were defined as eating meat, processed meat and poultry less than once a week. Other participants reporting to consume either meat or processed meat or poultry more often were categorized as non-vegetarians (9).

### Sample collection and handling

Native stool samples were collected by the participants at home from a normal bowel movement prior to bowel preparation for colonoscopy with standard stool collection tubes provided with a small spoon for collecting the stool. The stool samples were then directly frozen by the participants at -20°C at home. The participants were asked to document date and time of sampling and the storage temperature. The stool samples were taken by the participant in a freeze-cool transport container and in an isolated envelope to the gastroenterologists’ practices, directly frozen again at -20°C and within the week of receipt delivered by a transport service on dry ice to the GEKKO study laboratory at the National Center for Tumor Diseases (NCT) in Heidelberg where they were immediately frozen at -80°C.

### Processing of the samples

For this analysis, we used stool samples from 3 individuals of the GEKKO study testing 8 different stool processing protocols each in triplicate which results in a total of 72 measurements. Every protocol was performed with a frozen biomass of 50mg wet stool weight. Used protocols were adapted in part from previously published work (protocol 1 (10), protocol 3 (11) and protocol 7 (12)) and from the current recommended SOP provided from Biocrates (protocol 5 (13)). Details of the protocols are shown in **Table 1**. In brief, feces was cut and weighed in frozen state and kept frozen until processing. The samples were thawed and prepared according to the specific protocol and frozen again at -80°C until further processing via MxP® Quant 500 Kit (Biocrates Life Sciences AG, Innsbruck, Austria). Liquid sample extracts were processed according to the vendor’s instructions for human plasma samples. Samples dried after extraction had to be resolved before measurement (for protocol 6 and 7). Therefore, 50µl of 100% isopropanol were added into the vial and the mixture was vortexed for 3 min at room temperature. Additionally, 50µl of 30% isopropanol were added and again vortexed for 3min at room temperature. Short centrifugation (5sec) separated the solid substances from the liquid phase which was further used for analysis.

**Table 1.** Overview on the tested stool protocols.

| *Protocol 1* | *Protocol 2* | *Protocol 3* | *Protocol 4* | *Protocol 5* | *Protocol 6* | *Protocol 7* | *Protocol 8* |
| --- | --- | --- | --- | --- | --- | --- | --- |
| 50 mg stool | 50 mg stool | 50 mg stool | 50 mg stool | 50 mg stool | 50 mg stool | 50 mg stool | 50 mg stool |
| 150µL PBS | 200µl isopropanol | 200µl Methanol/<br>acetonitril/ H<sub>2</sub>O <sup>1</sup><br>(2/2/1) | 200µl Methanol/<br>Acetonitril (1/1) | 150µl 85% Ethanol/<br>15% 20mM<br>phosphate buffer | 200µl 75% Ethanol | 225µl 100%<br>Methanol | 150µl 100%<br>methanol |
| vortex 2 min |  |  |  |  |  |  |  |
|  |  |  |  |  |  | Freeze in liquid N<sub>2</sub><br>(1min); thaw |  |
| 5 min sonification on ice |  |  |  |  |  |  |  |
|  |  |  |  |  | 500µl MTBE | 750µl MTBE |  |
|  |  |  |  |  | shake 1 h at RT | shake 1h at 4°C |  |
|  |  |  |  |  |  | 188µl H<sub>2</sub>O <sup>1</sup> + 0.1%<br>ammonium acetate |  |
|  |  |  |  |  |  | vortex 2 min,<br>incubate 5 min at RT |  |
|  |  |  |  | Centrifugation at<br>full speed (5 min) |  | Centrifugation at full<br>speed (10 min) | Centrifugation<br>at full speed (5<br>min) |
|  |  |  |  | supernatant |  | Upper supernatant | supernatant |
|  |  |  |  | 150µl 20% Ethanol/<br>80% 20mM<br>phosphate buffer | 125µl H<sub>2</sub>O <sup>1</sup> | ~2000µl 100%<br>methanol | 150µl 20%<br>methanol |
|  |  |  |  |  | vortex 2 min | vortex 1 min |  |
|  |  |  |  |  | incubate 10 min at RT | Incubate 1h at -20°C |  |
| Centrifugation at full speed at 4°C (15 min) + supernatant in extra tube |  |  |  |  |  |  |  |
| Freeze in liquid<br>nitrogen | Freeze in liquid<br>nitrogen | Freeze in liquid<br>nitrogen | Freeze in liquid<br>nitrogen | Freeze in liquid<br>nitrogen | Dry complete in<br>SpeedVac | Dry complete in<br>SpeedVac | Freeze in liquid<br>nitrogen |

<sup>1</sup> DNase and RNase free water
**Abbreviations:** MTBE, methyl tert-butyl ether; PBS, phosphate buffered saline; RT, room temperature.

For metabolite measurements, a QTRAP6500+ (Sciex, Germany) MS/MS connected to an UPLC I-class Plus (Waters, Germany) chromatography system was used. Conditions for LC separation and FIA analyses as well as individual MRM parameters for each metabolite and respective internal standards were provided by the vendor of the kit (Biocrates). The software MetIDQ (version Oxygen; Biocrates Life Sciences AG, Innsbruck, Austria) was used for processing of the data.

In total, 630 metabolites can be measured via this kit. The limit of detection (LOD) for each compound is defined as three times the background noise. The lower limit of quantitation (LLOQ) is at least ten times the background noise. At LLOQ measured metabolite concentrations can be considered as reliable. Data are normalized with a tissue factor for quantification under the following assumption: 1mg tissue equals 1µl tissue or stool. Concentrations are given in pmol of the metabolite /mg stool mass.

The following metabolites are measurable using the aforementioned kit: 1 alkaloid (trigonelline), 1 amine oxide (trimethylamine N-oxide), 20 amino acids, 30 other amino acid related metabolites, 14 bile acids, 9 biogenic amines, sugars (hexoses including glucose), 7 carboxylic acids, 1 cresol (p-cresol sulfate), 12 fatty acids, 4 hormone and related metabolites, 4 indoles and derivatives, 2 nucleobases and related molecules, 1 metabolite from the group of vitamins and cofactors, 40 acylcarnitines, 14 lysophosphatidylcholines, 76 phosphatidylcholines, 15 sphingomyelins, 28 ceramides, 8 dihydroceramides, 19 hexosylceramides, 9 dihexosylceramides, 6 trihexosylceramides, 22 cholesteryl esters, 44 diglycerides, and 242 triglycerides. All the related isobaric and isomeric lipid species can be measured but cannot be distinguished by this method.

### Statistical analyses

We measured 630 metabolites and calculated a range of sums and ratios of metabolites indicating metabolic pathways and syntheses. Those compounds with the mean below LOD were excluded and described as not measured. Metabolism indicators were calculated with the MetIDQ software and those with more than half of the values below LOD were marked as below LOD in the following. We calculated means and standard deviations and assessed the number of metabolites and their respective classes for each processing method.

We described the study population that was used to apply the best protocol and used ANOVA to detect differences in metabolite concentrations between people with different dietary habits (vegetarians, semi-vegetarians, non-vegetarians).

A p-value <0.05 (two-sided testing) was considered to indicate statistical significance. Statistical analyses were conducted using SAS Enterprise Guide 7.1 (SAS Institute Inc., Cary, NC, USA).

## Results

We measured metabolites with the MxP® Quant 500 kit using 8 different protocols for stool processing from 3 healthy participants of the GEKKO study (free of neoplasms). We were able to extract metabolites with each protocol but the number of detectable metabolites varied (**Table 2**). Most metabolites were extracted (a) using isopropanol (protocol 2) and (b) when we dried the liquid extracts after extraction and reconstituted the samples in a smaller volume to increase metabolite concentrations (protocols 6 and 7).

**Table 2.** Number of metabolites that were measured with each protocol according to chemical class.

| Protocol | Total | 1 | 2 | 3 | 4 | 5 | 6 | 7 | 8 |
| --- | --- | --- | --- | --- | --- | --- | --- | --- | --- |
| analyte class |  |  |  |  |  |  |  |  |  |
| Total | 630 | 131 | 251 | 100 | 149 | 132 | 303 | 342 | 137 |
| Alkaloids | 1 | 1 | 1 | 1 | 1 | 1 | 1 | 1 | 1 |
| Amine Oxides | 1 | 0 | 0 | 0 | 0 | 0 | 0 | 0 | 0 |
| Amino Acids | 20 | 20 | 19 | 19 | 19 | 19 | 20 | 20 | 19 |
| Amino acid related | 30 | 25 | 22 | 23 | 23 | 22 | 26 | 25 | 23 |
| Bile Acids | 14 | 11 | 14 | 14 | 14 | 14 | 14 | 14 | 14 |
| Biogenic Amines | 9 | 8 | 6 | 6 | 6 | 6 | 7 | 7 | 6 |
| Carbohydrates and related | 1 | 1 | 0 | 0 | 0 | 0 | 1 | 0 | 0 |
| Carboxylic Acids | 7 | 4 | 2 | 3 | 2 | 2 | 3 | 4 | 2 |
| Cresols | 1 | 1 | 1 | 1 | 1 | 1 | 1 | 1 | 1 |
| Fatty Acids | 12 | 3 | 9 | 7 | 9 | 7 | 10 | 10 | 7 |
| Hormones and related | 4 | 1 | 2 | 2 | 2 | 1 | 2 | 2 | 2 |
| Indoles and Derivatives | 4 | 4 | 3 | 4 | 4 | 2 | 4 | 3 | 4 |
| Nucleobases and related | 2 | 2 | 2 | 2 | 2 | 2 | 2 | 2 | 2 |
| Vitamins and Cofactors | 1 | 1 | 1 | 1 | 1 | 1 | 1 | 1 | 1 |
| Acylcarnitines | 40 | 6 | 2 | 2 | 2 | 2 | 4 | 6 | 2 |
| Glycerophospholipide (Lysophosphatidylcholines and Phosphatidylcholines) | 90 | 5 | 25 | 7 | 18 | 10 | 27 | 38 | 10 |
| Sphingomyelins | 15 | 3 | 5 | 1 | 3 | 3 | 6 | 7 | 3 |
| Cholesteryl Esters | 22 | 3 | 3 | 0 | 0 | 0 | 4 | 6 | 0 |
| Ceramides | 28 | 0 | 22 | 0 | 15 | 10 | 24 | 26 | 6 |
| Dihydroceramides | 8 | 0 | 2 | 0 | 0 | 0 | 1 | 2 | 0 |
| Glycosylceramides(Mono-, Di-, and Trihexosylceramides) | 34 | 1 | 17 | 0 | 7 | 7 | 24 | 22 | 4 |
| Diglycerides | 44 | 6 | 14 | 4 | 11 | 5 | 17 | 18 | 4 |
| Triglycerides | 242 | 25 | 79 | 3 | 9 | 17 | 104 | 127 | 26 |
| Metabolism indicators | 232 | 66 | 72 | 85 | 78 | 86 | 99 | 96 | 84 |

The solvents differ in their polarity and therefore in their extraction efficacy to solve metabolites of the different chemical classes studied. With the MxP® Quant 500 kit, 630 metabolites can be determined of 14 classes of small molecules and 12 lipid classes (**Supplementary Table 1**). In addition, a range of sum and ratios that describe certain pathways and syntheses in the organism is calculated via the MetIDQ software from the obtained data (**Supplementary Table 2**). Amino acids and amino acid related products were detected with concentrations above limit of detection in the analyzed stool samples by all evaluated extraction protocols. Concentrations for amine oxides or carbohydrates and related products were always below the LOD. Major differences were observed for triglycerides as none of the compounds were above the LOD using PBS for preparation whereas more than 50 could be measured using isopropanol or ethanol or methanol in combination with concentrating the liquid extract.

Protocol 6 was favorable in terms of sample handling and measured numbers of metabolites. Therefore, we further measured 18 stool samples according to this protocol from participants with different dietary habits (**Table 3**). We analyzed stool samples from 18 vegetarians, semi-vegetarians and non-vegetarians and found a range of metabolites that were significantly different (**Table 4**). Most of the metabolites that distinguished the dietary habits were from lipid classes such as ceramides and phosphatidylcholines. Some metabolism indicators were also found to be different between vegetarians, semi-vegetarians and non-vegetarians. Most of the statistical significant ceramides were higher abundant in non-vegetarians and the sum of ceramides was increasing from vegetarians to non-vegetarians.

**Table 3.** Population characteristics for the GEKKO participants analyzed with protocol 6 by dietary habits.

| **Characteristics** | Vegetarians<br>N=6 | Semi-vegetarians<br>N=6 | Non-vegetarians<br>N=6 |
| --- | --- | --- | --- |
| **Sex, n (%)<sup>2</sup>** |  |  |  |
| Female | 3 | 3 | 3 |
| Male | 3 | 3 | 3 |
| **Age, n (%)<sup>2</sup>** |  |  |  |
| 50-54 years | 1 | 2 | 2 |
| 55-59 years | 4 | 2 | 4 |
| 60-64 years | 1 | 2 | 0 |
| Mean, (SD) | 56.7 ($\pm 2.9$) | 56.5 ($\pm 3.6$) | 54.8 ($\pm 2.6$) |

**Table 4.** Metabolite concentrations (and SD) and differences between vegetarians, semi-vegetarians and non-vegetarians.

| Metabolite | Class | vegetarians |  | semi-vegetarians |  | non-vegetarians |  | ANOVA<br>p value |
| --- | --- | --- | --- | --- | --- | --- | --- | --- |
|  |  | Mean | SD | Mean | SD | Mean | SD |  |
| Sum of Cer | Metabolism Indicators | 9.05 | 7.85 | 15.44 | 4.88 | 26.67 | 7.49 | 0.0017 |
| Cer(d16:1/23:0) | Ceramides | 0.18 | 0.21 | 0.34 | 0.06 | 0.56 | 0.14 | 0.0019 |
| Sum of LCFA-Cer | Metabolism Indicators | 4.82 | 4.69 | 8.62 | 2.57 | 16.16 | 5.74 | 0.0019 |
| Cer(d18:2/18:0) | Ceramides | 0.01 | 0.03 | 0.12 | 0.07 | 0.30 | 0.19 | 0.0028 |
| Cer(d16:1/22:0) | Ceramides | 0.37 | 0.32 | 0.53 | 0.15 | 0.96 | 0.25 | 0.0031 |
| Cer(d18:1/18:1) | Ceramides | 0.23 | 0.17 | 0.16 | 0.07 | 1.06 | 0.73 | 0.0046 |
| Cer(d18:0/22:0) | Dihydroceramides | 0.14 | 0.20 | 0.15 | 0.17 | 0.57 | 0.26 | 0.0049 |
| 1-Met-His Synthesis | Metabolism Indicators | 6.29 | 3.76 | 0.83 | 1.37 | 0.39 | 0.31 | 0.0058 |
| Ratio of Short-Chain to Long-Chain ACs | Metabolism Indicators | 0.95 | 0.32 | 0.80 | 0.29 | 0.46 | 0.07 | 0.0058 |
| Cer(d18:2/24:0) | Ceramides | 0.04 | 0.06 | 0.10 | 0.11 | 0.21 | 0.05 | 0.0061 |
| Sum of VLCFA-Cer | Metabolism Indicators | 4.24 | 3.32 | 6.81 | 2.66 | 10.53 | 2.69 | 0.0067 |
| PC ae C36:5 | Phosphatidylcholines | 0.03 | 0.04 | 0.06 | 0.05 | 0.21 | 0.14 | 0.0071 |
| Sum of VLCFA-DH-Cer | Metabolism Indicators | 0.65 | 0.24 | 1.11 | 0.55 | 2.35 | 0.95 | 0.0075 |
| SM C26:0 | Sphingomyelins | 0.02 | 0.02 | 0.01 | 0.01 | 0.05 | 0.02 | 0.0080 |
| PEA | Biogenic Amines | 0.15 | 0.14 | 0.18 | 0.19 | 0.97 | 0.74 | 0.0096 |
| CE(20:1) | Cholesteryl Esters | 1.14 | 0.71 | 0.12 | 0.30 | 0.22 | 0.54 | 0.0098 |
| Cer(d18:1/22:0) | Ceramides | 0.56 | 0.37 | 0.80 | 0.34 | 1.30 | 0.41 | 0.0115 |
| CE(14:1) | Cholesteryl Esters | 0.07 | 0.17 | 0.00 | 0.00 | 0.44 | 0.39 | 0.0147 |
| PEA Synthesis | Metabolism Indicators | 0.00 | 0.00 | 0.00 | 0.00 | 0.01 | 0.01 | 0.0157 |
| Cer(d18:0/24:1) | Dihydroceramides | 0.18 | 0.23 | 0.49 | 0.23 | 0.73 | 0.36 | 0.0157 |
| TG(17:1_32:1) | Triacylglycerides | 0.00 | 0.00 | 0.05 | 0.12 | 0.23 | 0.18 | 0.0158 |
| HexCer(d18:1/24:1) | Hexosylceramides | 2.45 | 1.16 | 5.48 | 2.63 | 7.24 | 3.29 | 0.0158 |
| Cer(d16:1/24:0) | Ceramides | 0.13 | 0.23 | 0.25 | 0.10 | 0.43 | 0.11 | 0.0178 |
| Sum of VLCFA-Glycosyl-Cer | Metabolism Indicators | 6.54 | 3.48 | 14.91 | 6.92 | 18.24 | 8.21 | 0.0200 |
| Cer(d18:1/24:1) | Ceramides | 0.89 | 0.72 | 1.31 | 0.59 | 2.12 | 0.71 | 0.0200 |
| Cer(d18:1/18:0(OH)) | Ceramides | 0.10 | 0.18 | 0.19 | 0.24 | 0.52 | 0.29 | 0.0213 |
| Sum of Glycosyl-Cer | Metabolism Indicators | 9.94 | 5.55 | 21.58 | 9.26 | 26.38 | 11.98 | 0.0224 |
| Sum of HexCer | Metabolism Indicators | 7.26 | 4.18 | 16.20 | 7.72 | 20.00 | 8.90 | 0.0227 |
| HexCer(d18:1/26:0) | Hexosylceramides | 0.21 | 0.14 | 0.41 | 0.17 | 0.52 | 0.22 | 0.0291 |
| TG(18:0_32:0) | Triacylglycerides | 1.79 | 0.21 | 1.46 | 0.27 | 1.34 | 0.32 | 0.0302 |
| Cer(d18:2/16:0) | Ceramides | 0.17 | 0.09 | 0.28 | 0.15 | 0.39 | 0.15 | 0.0308 |
| DG(17:0_18:1) | Diglycerides | 1.59 | 0.76 | 1.37 | 1.20 | 2.93 | 0.96 | 0.0316 |
| Cer(d18:1/20:0) | Ceramides | 0.11 | 0.08 | 0.10 | 0.06 | 0.21 | 0.08 | 0.0353 |
| C12-DC | Acylcarnitines | 1.20 | 0.36 | 0.90 | 0.15 | 1.42 | 0.37 | 0.0355 |
| PC ae C38:6 | Phosphatidylcholines | 0.03 | 0.02 | 0.07 | 0.04 | 0.14 | 0.11 | 0.0365 |
| Sum of LCFA-Glycosyl-Cer | Metabolism Indicators | 3.36 | 2.16 | 6.53 | 2.47 | 8.05 | 3.74 | 0.0366 |
| Cer(d18:1/24:0) | Ceramides | 0.54 | 0.44 | 0.97 | 0.52 | 1.37 | 0.52 | 0.0368 |
| Cer(d18:1/18:0) | Ceramides | 0.38 | 0.27 | 0.55 | 0.31 | 2.76 | 2.76 | 0.0378 |
| TG(18:1_32:2) | Triacylglycerides | 1.37 | 1.49 | 3.03 | 1.71 | 3.58 | 0.78 | 0.0381 |
| PC aa C34:4 | Phosphatidylcholines | 0.07 | 0.01 | 0.03 | 0.03 | 0.04 | 0.04 | 0.0395 |
| Cer(d18:2/18:1) | Ceramides | 0.03 | 0.02 | 0.04 | 0.02 | 0.09 | 0.05 | 0.0401 |
| PC ae C38:5 | Phosphatidylcholines | 0.07 | 0.03 | 0.13 | 0.06 | 0.17 | 0.08 | 0.0405 |
| PC ae C34:3 | Phosphatidylcholines | 0.02 | 0.04 | 0.18 | 0.20 | 0.54 | 0.53 | 0.0438 |
| CE(18:0) | Cholesteryl Esters | 1.13 | 0.57 | 0.92 | 0.41 | 3.02 | 2.41 | 0.0455 |
| Cer(d18:1/25:0) | Ceramides | 0.44 | 0.30 | 0.75 | 0.48 | 1.16 | 0.55 | 0.0472 |
| PC ae C44:4 | Phosphatidylcholines | 0.11 | 0.01 | 0.12 | 0.01 | 0.13 | 0.02 | 0.0498 |

**Abbreviations:** C12-DC, Dodecanedioylcarnitine; CE, Cholesteryl ester; Cer, Ceramide, DG, Diglyceride; LCFA, Long-chain fatty acid; PC, Phosphatidylcholine; PEA, Phenylethylamine; SM, Sphingomyelin; TG, Triglyceride.

## Discussion

There is no consensus so far how stool samples should be prepared for comparable, standardized metabolomics measurements (8, 14). We were able to extract a broad range of metabolites with each of the 8 defined protocols tested within this study. However, some methods should be preferred over others in regards to the solvents used dependent on the aim of the study and if a broad or a specific metabolic coverage is aimed for in a particular stool sample analysis. In this study the largest numbers of metabolites could be measured after extraction with isopropanol and ethanol or methanol following a drying step. We have seen differences in the measured metabolite concentration dependent on the extraction method.

We observed that the stool processing methods differ, are not interchangeable and that metabolite extraction efficiency varies. The metabolomics panels found by various research groups looking into metabolomics stool sample analysis differ in metabolite composition which might by caused, amongst other reasons, by the different stool processing methods. Studies that have focused on metabolomics in stool samples used either PBS/D_2_O buffer (6, 7), acetonitrile (5), methanol (15) or methanol/water mixture (1) for metabolite extraction and each buffer/ solvent results in specific biomarker panel for the specific reagent applied. In this analysis, we used PBS, acetonitrile, methanol and additionally other more complex biphasic (polar/ apolar) solvent combinations for metabolite extraction. The principal procedure is similar for all protocols. A solvent or mixture of solvents is added to the thawed stool samples and this mixture is homogenized and centrifuged so that that supernatant can be used for metabolite analysis (14). We were able to extract a broad range of metabolites with each procedure but the concentrations and the type of metabolites extracted, differ.

A total of 630 metabolites and various sums and ratios can be measured or calculated. The typical number of metabolites that can be measured with this kit in human stool is 117 using an ethanol phosphate buffer based protocol (16). When we used a protocol based on PBS, we were able to measure only 88 compounds. Using different methods, we were able to extract and quantify up to 340 metabolites from human stool samples. Amino acids, amino acid related metabolites, bile acids, fatty acids, nucleobases and related metabolites amongst others can be reliably measured with almost all solvents. In contrast, the numbers of acylcarnitines, glycerophospholipids or triglycerides are low in human stool which was also found by Wolf et al. (16).

In agreement with previously reported differences between fecal metabolomics of omnivores, vegetarians and vegans (17), we observed metabolite differences between vegetarians and non-vegetarians. In particular, we found major differences between stool samples from vegetarians and non-vegetarians in the lipid classes. Non-vegetarians were shown to have higher intake of total fat compared to vegetarians (18). The significant differences in amount of metabolites of lipid classes in non-vegetarians compared to vegetarians found in our study might reflect the difference in fat intake. In meat-eaters, higher blood levels of glycerophospholipids or sphingolipids were found compared to vegetarians or vegans as the most important sources for those metabolites are animal products (19). Furthermore, it was found by different studies that meat intake is associated with the TMAO metabolism as meat and meat products are rich in substances needed for the synthesis of TMAO (20). In contrast, we did not find any differences in fecal TMAO. Other studies found higher amounts of amino acid metabolites and bile acids excretion in urine from meat eaters as they have higher intake of proteins compared to people with high vegetable intake (21). In stool samples, we did not find different amounts of amino acids or bile acids between vegetarians and non-vegetarians.

There are some limitations of this study. First, various solvents have different abilities to dissolve and extract the metabolites of different classes and therefore there will be no comparable results when using different processing methods within one study or across several studies. There is a very broad range of chemical classes in metabolomics and not all metabolites can be extracted equally well with the same methods. Second, stool composition varies greatly depending on antibiotic use (22), diet or the water content that can be in a range from 63 to 85% (14). Drying the original stool samples taken out of the freezer before processing offers the advantage of referring to similar actual weights, avoiding bias due to difference in the water content among different samples (23). It is difficult though to imagine study participants to dry their stool samples and then freeze them in small amounts with equal weights for standardization and avoiding freeze-thaw cycles. One would also need to collect more wet weight stool to get enough dry weight stool for a standardized metabolomics analyses. In addition, it is still an obstacle for many people to collect their stool samples for study purposes, freeze them at home and finally transport them in a freezing device to their physician. And even for laboratories, processing stool samples in a consistent way is a challenge also due to its varying consistency. Another point are contaminations and variations introduced into the stool samples that can result from toilet water or from urine, which we tried to minimize using a stool collection aid (24). Further limitations are the small samples size and inclusion of healthy individuals only.

A major strength of this study is that we have tested various metabolite extraction protocols on the same analytical platform. Stool samples are very promising in metabolomics research for CRC as they directly represent the microbial activities and the cellular environment in the gut (4). The stool samples were only frozen once which should ensure good metabolite stability: Composition might locally differ since the 50mg were cut off as frozen biomass from the total stool sample without thawing and mixing the complete stool sample.

In conclusion, we found a broad range of metabolites measurable in human stool samples. Some chemical classes can be measured equally well with all protocols whereas others are highly dependent on the extraction method. The extraction methods using (a) isopropanol or (b) ethanol or methanol and MTBE including drying of the supernatant seem to be preferable over others for further metabolomics analyses. To our knowledge this is the first study in stool metabolomics comparing 8 different protocols for metabolite extraction with a novel highly standardized and quality controlled, quantitative and reproducible assay and evaluating one methodology, the for our purpose most favorable protocol, to evaluate metabolites in stool samples of participants with different dietary habits.

There is urgent need for a consensus on standard procedures for stool processing for metabolomics and for quantitative and reproducible assays to get comparable results across different studies and laboratories.

## Data Availability

none

## Abbreviations

CRC: colorectal cancer
MTBE: methyl tert-butyl ether

## Acknowledgement

The authors’ responsibilities were as follows – HB: planned and designed the GEKKO study; VE, PSK, HB: designed this specific analysis; GP: designed the protocols, VE, GP: processed the samples; HB, PSK: conducted the study; VE: analyzed the data; VE, PSK, HB: drafted the manuscript; all authors critically reviewed the manuscript and approved the final draft. The authors disclose no potential conflict of interests. We received the MxP® Quant 500 Kit (Biocrates Life Sciences AG, Innsbruck, Austria) used in this study free of charge and thank Biocrates for their kind support.

## References

1. Phua LC, Chue XP, Koh PK, Cheah PY, Ho HK, Chan EC. Non-invasive fecal metabonomic detection of colorectal cancer. Cancer biology & therapy 2014;15(4):389–97. doi: 10.4161/cbt.27625.

2. Rose C, Parker A, Jefferson B, Cartmell E. The Characterization of Feces and Urine: A Review of the Literature to Inform Advanced Treatment Technology. Crit Rev Environ Sci Technol 2015;45(17):1827–79. doi: 10.1080/10643389.2014.1000761.

3. Zhgun ES, Ilina EN. Fecal Metabolites As Non-Invasive Biomarkers of Gut Diseases. Acta naturae 2020;12(2):4–14. doi: 10.32607/actanaturae.10954.

4. Chetwynd AJ, Ogilvie LA, Nzakizwanayo J, Pazdirek F, Hoch J, Dedi C, Gilbert D, Abdul-Sada A, Jones BV, Hill EM. The potential of nanoflow liquid chromatography-nano electrospray ionisation-mass spectrometry for global profiling the faecal metabolome. Journal of chromatography A 2019;1600:127–36. doi: 10.1016/j.chroma.2019.04.028.

5. Amiot A, Dona AC, Wijeyesekera A, Tournigand C, Baumgaertner I, Lebaleur Y, Sobhani I, Holmes E. (1)H NMR Spectroscopy of Fecal Extracts Enables Detection of Advanced Colorectal Neoplasia. Journal of proteome research 2015;14(9):3871–81. doi: 10.1021/acs.jproteome.5b00277.

6. Bezabeh T, Somorjai R, Dolenko B, Bryskina N, Levin B, Bernstein CN, Jeyarajah E, Steinhart AH, Rubin DT, Smith IC. Detecting colorectal cancer by 1H magnetic resonance spectroscopy of fecal extracts. NMR in biomedicine 2009;22(6):593–600. doi: 10.1002/nbm.1372.

7. Lin Y, Ma C, Liu C, Wang Z, Yang J, Liu X, Shen Z, Wu R. NMR-based fecal metabolomics fingerprinting as predictors of earlier diagnosis in patients with colorectal cancer. Oncotarget 2016;7(20):29454–64. doi: 10.18632/oncotarget.8762.

8. Deda O, Gika HG, Wilson ID, Theodoridis GA. An overview of fecal sample preparation for global metabolic profiling. Journal of pharmaceutical and biomedical analysis 2015;113:137–50. doi: 10.1016/j.jpba.2015.02.006.

9. Orlich MJ, Singh PN, Sabate J, Fan J, Sveen L, Bennett H, Knutsen SF, Beeson WL, Jaceldo-Siegl K, Butler TL, et al. Vegetarian dietary patterns and the risk of colorectal cancers. JAMA internal medicine 2015;175(5):767–76. doi: 10.1001/jamainternmed.2015.59.

10. Bjerrum JT, Wang Y, Hao F, Coskun M, Ludwig C, Günther U, Nielsen OH. Metabonomics of human fecal extracts characterize ulcerative colitis, Crohn’s disease and healthy individuals. Metabolomics 2015;11:122–33. doi: 10.1007/s11306-014-0677-3.

11. Ivanisevic J, Zhu ZJ, Plate L, Tautenhahn R, Chen S, O’Brien PJ, Johnson CH, Marletta MA, Patti GJ, Siuzdak G. Toward ‘omic scale metabolite profiling: a dual separation-mass spectrometry approach for coverage of lipid and central carbon metabolism. Analytical chemistry 2013;85(14):6876–84. doi: 10.1021/ac401140h.

12. Lee DY, Kind T, Yoon YR, Fiehn O, Liu KH. Comparative evaluation of extraction methods for simultaneous mass-spectrometric analysis of complex lipids and primary metabolites from human blood plasma. Analytical and bioanalytical chemistry 2014;406(28):7275–86. doi: 10.1007/s00216-014-8124-x.

13. Biocrates Life Sciences AG. Analysis of Human Fecal Samples with the MxP(R) Quant 500 Kit. 2019.

14. Karu N, Deng L, Slae M, Guo AC, Sajed T, Huynh H, Wine E, Wishart DS. A review on human fecal metabolomics: Methods, applications and the human fecal metabolome database. Analytica chimica acta 2018;1030:1–24. doi: 10.1016/j.aca.2018.05.031.

15. Goedert JJ, Sampson JN, Moore SC, Xiao Q, Xiong X, Hayes RB, Ahn J, Shi J, Sinha R. Fecal metabolomics: assay performance and association with colorectal cancer. Carcinogenesis 2014;35(9):2089–96. doi: 10.1093/carcin/bgu131.

16. Wolf B, Heischmann S, Dearth S, Koal T. Spotlight. The MxP Quant 500 Kit. MetaboNews 2019:4–9.

17. De Filippis F, Pellegrini N, Vannini L, Jeffery IB, La Storia A, Laghi L, Serrazanetti DI, Di Cagno R, Ferrocino I, Lazzi C, et al. High-level adherence to a Mediterranean diet beneficially impacts the gut microbiota and associated metabolome. Gut 2016;65(11):1812–21. doi: 10.1136/gutjnl-2015-309957.

18. Rizzo NS, Jaceldo-Siegl K, Sabate J, Fraser GE. Nutrient profiles of vegetarian and nonvegetarian dietary patterns. J Acad Nutr Diet 2013;113(12):1610–9. doi: 10.1016/j.jand.2013.06.349.

19. Schmidt JA, Rinaldi S, Ferrari P, Carayol M, Achaintre D, Scalbert A, Cross AJ, Gunter MJ, Fensom GK, Appleby PN, et al. Metabolic profiles of male meat eaters, fish eaters, vegetarians, and vegans from the EPIC-Oxford cohort. Am J Clin Nutr 2015;102(6):1518–26. doi: 10.3945/ajcn.115.111989.

20. Guasch-Ferré M, Bhupathiraju SN, Hu FB. Use of Metabolomics in Improving Assessment of Dietary Intake. Clinical chemistry 2018;64(1):82–98. doi: 10.1373/clinchem.2017.272344.

21. Wei R, Ross AB, Su M, Wang J, Guiraud S-P, Draper CF, Beaumont M, Jia W, Martin F-P. Metabotypes Related to Meat and Vegetable Intake Reflect Microbial, Lipid and Amino Acid Metabolism in Healthy People. Molecular Nutrition & Food Research 2018;62(21):1800583. doi: 10.1002/mnfr.201800583.

22. Pérez-Cobas AE, Gosalbes MJ, Friedrichs A, Knecht H, Artacho A, Eismann K, Otto W, Rojo D, Bargiela R, von Bergen M, et al. Gut microbiota disturbance during antibiotic therapy: a multi-omic approach. Gut 2013;62(11):1591–601. doi: 10.1136/gutjnl-2012-303184.

23. Deda O, Chatziioannou AC, Fasoula S, Palachanis D, Raikos N, Theodoridis GA, Gika HG. Sample preparation optimization in fecal metabolic profiling. Journal of chromatography B, Analytical technologies in the biomedical and life sciences 2017;1047:115–23. doi: 10.1016/j.jchromb.2016.06.047.

24. Wu WK, Chen CC, Panyod S, Chen RA, Wu MS, Sheen LY, Chang SC. Optimization of fecal sample processing for microbiome study - The journey from bathroom to bench. Journal of the Formosan Medical Association = Taiwan yi zhi 2019;118(2):545–55. doi: 10.1016/j.jfma.2018.02.005.

