## Supplementary Tables for "Evaluation of different stool extraction methods for metabolomics measurements in human fecal samples"

**Supplementary Table 1**. Concentrations of metabolites (in pmol/mg stool) and their standard deviations that were measured in at least one of the tested protocols >LOD.

| **Protocol** | **1** | | **2** | | **3** | | **4** | | **5** | | **6** | | **7** | | **8** | |
|---|---|---|---|---|---|---|---|---|---|---|---|---|---|---|---|---|
| **Metabolite** | **mean** | **SD** | **mean** | **SD** | **mean** | **SD** | **mean** | **SD** | **mean** | **SD** | **mean** | **SD** | **mean** | **SD** | **mean** | **SD** |
| C0 | 47.62 | 21.67 | 17.38 | 11.66 | 31.74 | 17.59 | 27.22 | 14.29 | 39.34 | 27.32 | 24.5 | 13.97 | 26.23 | 14.5 | 42.19 | 24.98 |
| C7-DC | 0.53 | 0.28 | 0.23 | 0.06 | 0.34 | 0.23 | 0.3 | 0.13 | 0.41 | 0.15 | 0.15 | 0.04 | 0.23 | 0.16 | 0.38 | 0.23 |
| C9 | 0.45 | 0.29 | 0.12 | 0.01 | 0.14 | 0.02 | 0.14 | 0.02 | 0.21 | 0.04 | 0.09 | 0.01 | 0.1 | 0.02 | 0.2 | 0.03 |
| C10 | 1.39 | 0.88 | 0.5 | 0.06 | 0.52 | 0.1 | 0.51 | 0.07 | 0.74 | 0.14 | 0.36 | 0.05 | 0.4 | 0.08 | 0.71 | 0.12 |
| C12 | 0.82 | 0.43 | 0.34 | 0.06 | 0.36 | 0.09 | 0.36 | 0.09 | 0.45 | 0.08 | 0.31 | 0.29 | 0.23 | 0.06 | 0.46 | 0.09 |
| C12:1 | 0.88 | 0.48 | 0.41 | 0.07 | 0.51 | 0.09 | 0.44 | 0.08 | 0.65 | 0.09 | 0.3 | 0.08 | 0.47 | 0.16 | 0.64 | 0.19 |
| C16 | 0.18 | 0.03 | 0.37 | 0.21 | 0.38 | 0.16 | 0.38 | 0.22 | 0.39 | 0.15 | 0.41 | 0.18 | 0.45 | 0.26 | 0.44 | 0.14 |
| C16:2 | 0.08 | 0.01 | 0.09 | 0.02 | 0.12 | 0.03 | 0.12 | 0.03 | 0.13 | 0.02 | 0.12 | 0.02 | 0.1 | 0.02 | 0.14 | 0.03 |
| C18 | 0.09 | 0.02 | 0.64 | 0.41 | 0.37 | 0.18 | 0.55 | 0.34 | 0.44 | 0.22 | 0.4 | 0.23 | 0.59 | 0.35 | 0.49 | 0.27 |
| C18:1 | 0.1 | 0.02 | 0.26 | 0.1 | 0.28 | 0.1 | 0.31 | 0.13 | 0.3 | 0.11 | 0.24 | 0.11 | 0.3 | 0.17 | 0.34 | 0.07 |
| Trigonelline | 3.28 | 2.01 | 1.55 | 0.98 | 2.29 | 0.96 | 2.13 | 0.99 | 2.64 | 1.5 | 2.05 | 0.82 | 1.84 | 0.64 | 2.84 | 1.64 |
| Ala | 1041.78 | 237.98 | 239.89 | 82.87 | 471 | 166.58 | 345.11 | 125.77 | 509.56 | 86.92 | 1625.89 | 978.93 | 457 | 118.79 | 560.89 | 166.13 |
| Arg | 134.78 | 42.19 | 14.6 | 22.1 | 56.67 | 64.01 | 17.61 | 27.15 | 66.57 | 51.88 | 365 | 62.11 | 137.74 | 132.2 | 141.41 | 122.98 |
| Asn | 47.3 | 35.07 | 2.94 | 1.01 | 7.26 | 4.25 | 3.59 | 1.77 | 9.57 | 5.15 | 108.79 | 83.09 | 8.55 | 4.28 | 9.06 | 4.42 |
| Asp | 1223.11 | 350.03 | 79.14 | 55.31 | 652.89 | 475.11 | 207.3 | 189.99 | 873.44 | 315.86 | 881.33 | 476.9 | 453.11 | 182.05 | 1003.78 | 396.9 |
| Cys | 58.92 | 21.02 | 13.32 | 6.88 | 37.23 | 8.06 | 20.57 | 5.59 | 52.1 | 15.4 | 31.19 | 9.56 | 25.91 | 7.94 | 50.78 | 12.51 |
| Gln | 220.78 | 46.27 | 28.83 | 11.51 | 83.63 | 30.97 | 47.99 | 22.01 | 83.6 | 31.42 | 221.14 | 127.27 | 71.76 | 27.61 | 97.69 | 36.99 |
| Glu | 5207.11 | 2707.07 | 507.44 | 292.03 | 3912.22 | 3471.97 | 1946.44 | 2117.8 | 4124.33 | 2339.93 | 2814.44 | 1629.6 | 2561 | 1736.3 | 5053 | 3244.98 |
| Gly | 451.78 | 76.63 | 83.11 | 31.78 | 247.44 | 110.96 | 135.97 | 69.34 | 278.89 | 89.61 | 479.22 | 221.66 | 169.44 | 30.16 | 298.11 | 114.03 |
| His | 97.86 | 14.88 | 27.47 | 8.71 | 52.51 | 17.09 | 35.64 | 9.56 | 58.39 | 11.97 | 66.62 | 42.75 | 48.5 | 11.48 | 77.08 | 21.75 |
| Ile | 212.56 | 77.9 | 39.51 | 12 | 65.18 | 29.84 | 54.48 | 25.06 | 67.48 | 15.38 | 416.43 | 304.01 | 67.29 | 29.59 | 75.42 | 34.45 |
| Leu | 366.33 | 105.85 | 58.57 | 24.15 | 101.6 | 48.87 | 82.43 | 44.28 | 105.58 | 29.9 | 981 | 779.27 | 126.9 | 55.71 | 118.71 | 60.92 |
| Lys | 1069.89 | 203.46 | 169.36 | 122.28 | 526 | 156.8 | 235.26 | 145.84 | 700 | 201.59 | 878.11 | 404.28 | 400.89 | 216.22 | 805.89 | 172.15 |

| | | | | | | | | | [20] | | | | | | | |
|---|---|---|---|---|---|---|---|---|---|---|---|---|---|---|---|---|
| **Protocol** | **1** | | **2** | | **3** | | **4** | | **5** | | **6** | | **7** | | **8** | |
| **Metabolite** | **mean** | **SD** | **mean** | **SD** | **mean** | **SD** | **mean** | **SD** | **mean** | **SD** | **mean** | **SD** | **mean** | **SD** | **mean** | **SD** |
| Met | 197 | 54.94 | 33.49 | 12.78 | 63.98 | 33.77 | 47.97 | 29.5 | 61.91 | 21.56 | 379.46 | 257.98 | 50.92 | 24.1 | 64.07 | 30.86 |
| Phe | 187.89 | 45.47 | 29.64 | 10.87 | 52.23 | 23.29 | 42.34 | 20.34 | 52.5 | 14.58 | 382.56 | 254.64 | 55.74 | 23.53 | 62.41 | 28.22 |
| Pro | 349 | 108.32 | 139 | 60.06 | 259.78 | 164.07 | 216.94 | 129.55 | 247.77 | 117.23 | 285.44 | 128.85 | 197.72 | 98.81 | 312.24 | 165.21 |
| Ser | 298.78 | 96.61 | 35.3 | 11.37 | 101.03 | 44.07 | 57.7 | 31.31 | 108.43 | 28.91 | 384.56 | 215.11 | 64.51 | 24.33 | 111.3 | 47.58 |
| Thr | 200.22 | 43.81 | 38.76 | 12.23 | 87.37 | 30 | 57.17 | 26.32 | 87.17 | 19.5 | 265.21 | 165.08 | 75.9 | 27.34 | 96.93 | 28.54 |
| Trp | 44.2 | 14.57 | 9.54 | 7.28 | 18.79 | 12.13 | 14.17 | 10.31 | 15.27 | 7.45 | 56.61 | 32.23 | 16.65 | 8.67 | 20.01 | 10.68 |
| Tyr | 254.11 | 55.75 | 68.44 | 20.08 | 112.97 | 44.6 | 90.49 | 35.05 | 113.72 | 23.34 | 460.11 | 301.35 | 120.81 | 44.34 | 142.9 | 47.96 |
| Val | 312.22 | 71.32 | 81.7 | 35.19 | 133.56 | 37.22 | 112.07 | 31.8 | 147 | 46.62 | 540 | 319.3 | 143.11 | 44.12 | 160.29 | 53.14 |
| 1-Met-His | 3.92 | 4.1 | 1.53 | 1.65 | 2.77 | 2.32 | 2.08 | 1.75 | 3.6 | 3.47 | 2.35 | 2.11 | 2.18 | 1.68 | 4.03 | 4.25 |
| 3-Met-His | 9.96 | 3.83 | 3.62 | 2 | 7.06 | 2.02 | 5.23 | 1.98 | 8.92 | 4.3 | 6 | 2.58 | 6.51 | 2.63 | 9.27 | 3.24 |
| 5-AVA | 272 | 136.86 | 129.98 | 71.29 | 232.33 | 65.69 | 180.91 | 66.99 | 301.22 | 152.68 | 208 | 75.91 | 251.11 | 117.57 | 285.44 | 109.23 |
| AABA | 23.71 | 9.13 | 11.29 | 6.57 | 19.62 | 7.22 | 16.2 | 4.43 | 20.22 | 8.93 | 25.19 | 11.88 | 29.42 | 16.59 | 27.06 | 10.7 |
| Ac-Orn | 10.28 | 6.64 | 3.26 | 3.06 | 5.86 | 3.41 | 4.45 | 2.67 | 8.01 | 7.15 | 5.36 | 4.22 | 4.9 | 3.33 | 9.23 | 8.03 |
| ADMA | 1.18 | 0.49 | 0.48 | 0.3 | 0.93 | 0.16 | 0.72 | 0.21 | 1.15 | 0.42 | 0.8 | 0.26 | 0.85 | 0.22 | 1.27 | 0.38 |
| alpha-AAA | 5.04 | 1.88 | 0.88 | 0.51 | 3.55 | 0.96 | 1.68 | 0.73 | 4.88 | 2.14 | 3.18 | 1.52 | 3 | 1.34 | 4.89 | 1.82 |
| Anserine | 3.12 | 1.58 | 0.82 | 0.65 | 2.12 | 1.07 | 1.39 | 0.98 | 2.44 | 1.48 | 2.18 | 1.47 | 2.08 | 1.02 | 2.74 | 1.37 |
| BABA | 0.96 | 0.42 | 0.61 | 0.46 | 0.91 | 0.47 | 0.79 | 0.55 | 0.99 | 0.45 | 0.78 | 0.5 | 0.81 | 0.44 | 1.16 | 0.55 |
| Betaine | 0 | 0 | 1.63 | 2.61 | 1.82 | 2.79 | 1.16 | 2.35 | 0 | 0 | 1.35 | 2.08 | 2.67 | 4.34 | 2.11 | 3.25 |
| C4-OH-Pro | 1.65 | 1.41 | 1 | 0.5 | 1.33 | 0.76 | 1.05 | 0.49 | 1.83 | 1.33 | 0.93 | 0.62 | 0.95 | 0.51 | 1.85 | 1.21 |
| Carnosine | 2.97 | 2.38 | 1.25 | 0.45 | 2.14 | 1.19 | 1.42 | 0.37 | 2.96 | 1.98 | 2.37 | 1.85 | 2.25 | 1.53 | 3.1 | 2.13 |
| Cit | 503.56 | 150.98 | 105.32 | 38.69 | 269.78 | 121.54 | 164.41 | 85.01 | 326.67 | 107.72 | 650.56 | 587.23 | 164.61 | 55.7 | 279.78 | 94.54 |
| Creatinine | 10.66 | 4.07 | 14.07 | 9.14 | 16.95 | 11.58 | 15.79 | 10.84 | 20.57 | 12.28 | 13.75 | 10.42 | 20.63 | 21.15 | 18.16 | 11.08 |
| Cystine | 1.27 | 0.71 | 0.07 | 0.03 | 0.23 | 0.09 | 0.09 | 0.03 | 0.27 | 0.21 | 1.84 | 1.92 | 0.27 | 0.13 | 0.35 | 0.21 |
| DOPA | 0.66 | 0.31 | 0.47 | 0.18 | 0.65 | 0.25 | 0.56 | 0.21 | 0.84 | 0.36 | 0.49 | 0.17 | 0.4 | 0.17 | 0.8 | 0.33 |
| HArg | 0.66 | 0.14 | 0.21 | 0.12 | 0.46 | 0.21 | 0.26 | 0.15 | 0.55 | 0.24 | 0.52 | 0.27 | 0.4 | 0.25 | 0.63 | 0.21 |
| HCys | 15.14 | 1.52 | 10.82 | 3.55 | 12.66 | 4.19 | 10.65 | 3.73 | 16.53 | 4.54 | 16.06 | 9.74 | 12.36 | 8.26 | 16.38 | 3.62 |
| Met-SO | 10.93 | 5.94 | 3.51 | 1.33 | 5.94 | 3.86 | 4.62 | 2.79 | 4.96 | 1.44 | 23.32 | 13.47 | 11.32 | 7.53 | 5.34 | 3.63 |
| Orn | 116.02 | 76.18 | 18.96 | 19.61 | 42.64 | 29.8 | 17.03 | 10.93 | 70.34 | 60.48 | 50.69 | 27.53 | 24.56 | 19.72 | 80.04 | 68.39 |
| PheAlaBetaine | 1.02 | 1.37 | 0.68 | 0.94 | 0.68 | 0.87 | 0.76 | 1.08 | 0.74 | 0.94 | 0.61 | 0.81 | 0.59 | 0.76 | 0.8 | 1.1 |
| ProBetaine | 8.36 | 8.88 | 5.5 | 5.42 | 6.92 | 5.56 | 6.71 | 5.81 | 8.36 | 7.62 | 5.63 | 4.71 | 5.69 | 4 | 9.23 | 8.47 |

[21]

| Protocol | 1 | | 2 | | 3 | | 4 | | 5 | | 6 | | 7 | | 8 | |
|---|---|---|---|---|---|---|---|---|---|---|---|---|---|---|---|---|
| **Metabolite** | **mean** | **SD** | **mean** | **SD** | **mean** | **SD** | **mean** | **SD** | **mean** | **SD** | **mean** | **SD** | **mean** | **SD** | **mean** | **SD** |
| Sarcosine | 20.55 | 17.3 | 9.33 | 7.59 | 15.92 | 8.26 | 12.76 | 6.24 | 21.84 | 18.26 | 13.57 | 7.89 | 15.68 | 9.23 | 21.15 | 15.72 |
| SDMA | 0.71 | 0.2 | 0.37 | 0.18 | 0.66 | 0.2 | 0.49 | 0.2 | 0.79 | 0.28 | 0.53 | 0.18 | 0.66 | 0.34 | 0.82 | 0.2 |
| t4-OH-Pro | 25.94 | 22.97 | 11.7 | 10.37 | 17.91 | 10.55 | 13.56 | 7.48 | 22.16 | 18.68 | 16.3 | 12.51 | 15.33 | 9.35 | 24.13 | 19.53 |
| Taurine | 41.6 | 22.94 | 27.22 | 23.17 | 46.49 | 37.7 | 39.03 | 38.97 | 53.3 | 43.31 | 49.45 | 39.13 | 52.05 | 60.68 | 46.45 | 35.02 |
| TrpBetaine | 0.35 | 0.22 | 0.19 | 0.13 | 0.23 | 0.15 | 0.21 | 0.13 | 0.28 | 0.2 | 0.25 | 0.16 | 0.19 | 0.12 | 0.32 | 0.25 |
| CA | 10.32 | 14.3 | 15.69 | 18.75 | 22.25 | 25.97 | 15.67 | 14.73 | 21.55 | 23.65 | 23.44 | 31.35 | 19.47 | 17.42 | 18.54 | 21 |
| CDCA | 1.24 | 1.85 | 12.32 | 14.59 | 24.47 | 36.32 | 15.72 | 18.43 | 11.66 | 8.2 | 32.92 | 53.2 | 18.37 | 18.11 | 11.4 | 7.19 |
| DCA | 21.19 | 16.4 | 72.89 | 42.78 | 80.49 | 43.43 | 73.93 | 46.8 | 93.97 | 54.87 | 73.54 | 38.48 | 76.24 | 39.96 | 98.21 | 60.89 |
| GCA | 0.07 | 0.05 | 0.32 | 0.25 | 0.9 | 0.84 | 0.84 | 1.04 | 0.8 | 0.79 | 0.63 | 0.41 | 0.83 | 0.8 | 1.58 | 1.96 |
| GCDCA | 0.09 | 0.03 | 1.42 | 2.73 | 2.8 | 3.78 | 1.55 | 2.16 | 1.07 | 0.59 | 3.51 | 6.44 | 3.71 | 4.44 | 1.37 | 0.82 |
| GDCA | 0.01 | 0.01 | 0.35 | 0.67 | 0.65 | 1.23 | 0.42 | 0.81 | 0.22 | 0.17 | 0.93 | 1.87 | 0.87 | 1.57 | 0.24 | 0.17 |
| GLCA | 0.01 | 0 | 0.05 | 0.03 | 0.05 | 0.03 | 0.06 | 0.05 | 0.06 | 0.03 | 0.06 | 0.04 | 0.06 | 0.05 | 0.05 | 0.03 |
| GLCAS | 0.01 | 0.01 | 0.37 | 0.56 | 0.65 | 0.98 | 0.82 | 1.47 | 0.58 | 0.88 | 0.41 | 0.59 | 1.46 | 2.51 | 0.61 | 0.84 |
| GUDCA | 0.05 | 0.04 | 0.03 | 0.03 | 0.07 | 0.03 | 0.03 | 0.02 | 0.06 | 0.03 | 0.18 | 0.13 | 0.06 | 0.04 | 0.06 | 0.04 |
| TCA | 0.21 | 0.22 | 1.01 | 0.94 | 1.52 | 1.54 | 1.73 | 2.38 | 1.39 | 0.65 | 0.82 | 1.8 | 2.35 | 3.01 | 1.9 | 0.92 |
| TCDCA | 0.04 | 0.02 | 2.73 | 5.01 | 4.77 | 9.69 | 2.61 | 4.37 | 1.56 | 0.88 | 1.76 | 4.49 | 3.32 | 4.46 | 1.93 | 1.13 |
| TDCA | 0.02 | 0.01 | 0.46 | 0.82 | 1.1 | 2.58 | 0.61 | 1.31 | 0.26 | 0.21 | 0.6 | 1.59 | 0.88 | 1.86 | 0.29 | 0.25 |
| TLCA | 0.01 | 0.01 | 0.13 | 0.14 | 0.21 | 0.22 | 0.22 | 0.33 | 0.17 | 0.19 | 0.06 | 0.05 | 0.33 | 0.49 | 0.16 | 0.16 |
| TMCA | 0.07 | 0.08 | 0.06 | 0.08 | 0.06 | 0.07 | 0.05 | 0.06 | 0.08 | 0.1 | 0.05 | 0.07 | 0.06 | 0.06 | 0.08 | 0.11 |
| beta-Ala | 36.71 | 10.5 | 13.11 | 4.94 | 30.88 | 16.02 | 19.4 | 8.99 | 32.51 | 9.53 | 31.08 | 12.71 | 25.21 | 6.97 | 37.57 | 14.53 |
| GABA | 22.86 | 16.75 | 11.23 | 8.7 | 20.44 | 11.15 | 15.64 | 9.71 | 25.98 | 16.99 | 21.04 | 11.88 | 22.63 | 15.2 | 35.81 | 33.56 |
| Histamine | 5.21 | 3.92 | 5.41 | 4.65 | 6.19 | 5.56 | 5.91 | 5.43 | 7.38 | 6.26 | 5.31 | 5.22 | 5.41 | 5.72 | 6.95 | 5.2 |
| PEA | 0.01 | 0.01 | 0 | 0.01 | 0 | 0 | 0 | 0 | 0 | 0.01 | 0.04 | 0.03 | 0.03 | 0.02 | 0 | 0 |
| Putrescine | 42.87 | 21.7 | 19.56 | 13.97 | 27.13 | 15.6 | 18.27 | 11.85 | 33.46 | 19.69 | 23.6 | 8.17 | 15.47 | 8.57 | 31.27 | 20.56 |
| Serotonin | 0.62 | 0.29 | 0.65 | 0.29 | 0.78 | 0.27 | 0.74 | 0.24 | 0.85 | 0.36 | 0.59 | 0.23 | 0.6 | 0.17 | 0.94 | 0.43 |
| Spermidine | 153.33 | 30.87 | 37.36 | 20.28 | 40.09 | 21.64 | 25.76 | 21.27 | 45.78 | 13.97 | 37.71 | 13.83 | 21.48 | 14.72 | 40.33 | 10.89 |
| Spermine | 3.49 | 0.53 | 1.17 | 0.25 | 1.09 | 0.15 | 0.95 | 0.14 | 1.58 | 0.34 | 0.99 | 0.23 | 0.67 | 0.23 | 1.56 | 0.36 |
| AconAcid | 2.75 | 1.72 | 0.23 | 0.09 | 0.39 | 0.06 | 0.3 | 0.08 | 0.44 | 0.15 | 0.33 | 0.13 | 0.33 | 0.11 | 0.48 | 0.12 |
| DiCA(12:0) | 8.43 | 11.15 | 2.6 | 2.65 | 4.37 | 5.16 | 3.64 | 4.17 | 4.63 | 5.13 | 2.06 | 2.29 | 3.4 | 4.18 | 5.27 | 6.06 |
| DiCA(14:0) | 0.08 | 0.04 | 0.12 | 0.05 | 0.12 | 0.05 | 0.11 | 0.04 | 0.19 | 0.05 | 0.13 | 0.1 | 0.17 | 0.13 | 0.18 | 0.07 |

[22]

| Protocol | 1 | | 2 | | 3 | | 4 | | 5 | | 6 | | 7 | | 8 | |
|---|---|---|---|---|---|---|---|---|---|---|---|---|---|---|---|---|
| **Metabolite** | **mean** | **SD** | **mean** | **SD** | **mean** | **SD** | **mean** | **SD** | **mean** | **SD** | **mean** | **SD** | **mean** | **SD** | **mean** | **SD** |
| OH-GlutAcid | 48.71 | 43.5 | 0.79 | 0.22 | 12.53 | 20.21 | 1.98 | 1.6 | 4.57 | 2.53 | 5.91 | 9.84 | 4.54 | 6.63 | 4.88 | 2.86 |
| Suc | 1100.67 | 1047.79 | 94.82 | 102.69 | 398.78 | 211.61 | 195.21 | 84.3 | 602.78 | 489.02 | 384.83 | 229.87 | 508.89 | 316.29 | 601.11 | 447.87 |
| Cer(d16:1/18:0) | 0 | 0 | 0.16 | 0.17 | 0.02 | 0.04 | 0.09 | 0.12 | 0.02 | 0.06 | 0.16 | 0.08 | 0.17 | 0.13 | 0.03 | 0.09 |
| Cer(d16:1/20:0) | 0 | 0 | 0.08 | 0.07 | 0.02 | 0.04 | 0.04 | 0.08 | 0 | 0 | 0.05 | 0.05 | 0.06 | 0.06 | 0.04 | 0.07 |
| Cer(d16:1/22:0) | 0 | 0 | 0.63 | 0.32 | 0 | 0 | 0.18 | 0.23 | 0.12 | 0.14 | 0.35 | 0.17 | 0.51 | 0.25 | 0.1 | 0.15 |
| Cer(d16:1/23:0) | 0 | 0 | 0.54 | 0.27 | 0 | 0 | 0.07 | 0.14 | 0.08 | 0.13 | 0.33 | 0.09 | 0.41 | 0.16 | 0.07 | 0.13 |
| Cer(d16:1/24:0) | 0.01 | 0.03 | 0.3 | 0.15 | 0.01 | 0.03 | 0.06 | 0.13 | 0.04 | 0.09 | 0.13 | 0.12 | 0.22 | 0.16 | 0.02 | 0.06 |
| Cer(d18:1/14:0) | 0 | 0 | 0.26 | 0.07 | 0.04 | 0.05 | 0.16 | 0.13 | 0.09 | 0.13 | 0.24 | 0.07 | 0.2 | 0.18 | 0.09 | 0.11 |
| Cer(d18:1/16:0) | 0.06 | 0.07 | 1.81 | 0.99 | 0.03 | 0.06 | 0.68 | 0.49 | 0.56 | 0.24 | 1.03 | 0.27 | 1.49 | 0.7 | 0.6 | 0.35 |
| Cer(d18:1/18:0(OH)) | 0 | 0 | 0.14 | 0.15 | 0 | 0 | 0.12 | 0.18 | 0.04 | 0.11 | 0.15 | 0.16 | 0.26 | 0.17 | 0.05 | 0.15 |
| Cer(d18:1/18:0) | 0.03 | 0.06 | 2.16 | 1.42 | 0.01 | 0.03 | 0.49 | 0.44 | 0.5 | 0.39 | 0.79 | 0.31 | 1.29 | 0.73 | 0.45 | 0.31 |
| Cer(d18:1/18:1) | 0.05 | 0.05 | 0.72 | 0.32 | 0.08 | 0.07 | 0.47 | 0.24 | 0.36 | 0.08 | 0.58 | 0.15 | 0.72 | 0.19 | 0.42 | 0.17 |
| Cer(d18:1/20:0(OH)) | 0.14 | 0.43 | 3.5 | 2.81 | 0 | 0 | 0.71 | 1.41 | 0.66 | 1.33 | 2.32 | 1.87 | 4.39 | 3.3 | 0.66 | 1.32 |
| Cer(d18:1/20:0) | 0.01 | 0.01 | 0.24 | 0.14 | 0 | 0 | 0.06 | 0.1 | 0.04 | 0.08 | 0.12 | 0.04 | 0.19 | 0.09 | 0.05 | 0.06 |
| Cer(d18:1/22:0) | 0 | 0 | 1.29 | 0.71 | 0 | 0 | 0.19 | 0.23 | 0.2 | 0.18 | 0.56 | 0.15 | 0.79 | 0.36 | 0.1 | 0.12 |
| Cer(d18:1/23:0) | 0 | 0 | 1.15 | 0.51 | 0 | 0 | 0.11 | 0.19 | 0.28 | 0.13 | 0.47 | 0.15 | 0.66 | 0.3 | 0.09 | 0.15 |
| Cer(d18:1/24:0) | 0.05 | 0.09 | 1.55 | 0.85 | 0.02 | 0.05 | 0.24 | 0.5 | 0.23 | 0.25 | 0.61 | 0.21 | 0.83 | 0.43 | 0.13 | 0.11 |
| Cer(d18:1/24:1) | 0.04 | 0.06 | 1.65 | 0.74 | 0 | 0 | 0.25 | 0.22 | 0.33 | 0.28 | 0.9 | 0.25 | 1.31 | 0.61 | 0.14 | 0.14 |
| Cer(d18:1/25:0) | 0.01 | 0.04 | 0.71 | 0.38 | 0.05 | 0.08 | 0.18 | 0.19 | 0.26 | 0.22 | 0.53 | 0.1 | 0.69 | 0.27 | 0.09 | 0.15 |
| Cer(d18:1/26:0) | 0.01 | 0.02 | 0.08 | 0.06 | 0 | 0 | 0.05 | 0.12 | 0.02 | 0.04 | 0.04 | 0.03 | 0.09 | 0.11 | 0 | 0 |
| Cer(d18:2/16:0) | 0 | 0 | 0.18 | 0.1 | 0.02 | 0.04 | 0.14 | 0.12 | 0.15 | 0.1 | 0.18 | 0.06 | 0.19 | 0.05 | 0.13 | 0.08 |
| Cer(d18:2/18:0) | 0 | 0 | 0.12 | 0.1 | 0.01 | 0.03 | 0.08 | 0.1 | 0 | 0 | 0.1 | 0.06 | 0.12 | 0.08 | 0.03 | 0.08 |
| Cer(d18:2/18:1) | 0 | 0 | 0.03 | 0.01 | 0 | 0.01 | 0.02 | 0.02 | 0.01 | 0.02 | 0.03 | 0.01 | 0.03 | 0.02 | 0.01 | 0.02 |
| Cer(d18:2/20:0) | 0 | 0 | 0.01 | 0.02 | 0 | 0 | 0.01 | 0.03 | 0 | 0 | 0.02 | 0.04 | 0.05 | 0.06 | 0 | 0 |
| Cer(d18:2/22:0) | 0 | 0 | 0.16 | 0.15 | 0 | 0 | 0.04 | 0.09 | 0.02 | 0.05 | 0.06 | 0.09 | 0.19 | 0.11 | 0 | 0 |
| Cer(d18:2/23:0) | 0 | 0 | 0.02 | 0.04 | 0 | 0.01 | 0 | 0 | 0 | 0 | 0.05 | 0.05 | 0.04 | 0.05 | 0 | 0 |
| Cer(d18:2/24:0) | 0 | 0 | 0.11 | 0.14 | 0 | 0 | 0.04 | 0.09 | 0 | 0 | 0.06 | 0.11 | 0.15 | 0.13 | 0 | 0 |
| Cer(d18:2/24:1) | 0 | 0 | 0.17 | 0.13 | 0 | 0 | 0.02 | 0.06 | 0.02 | 0.04 | 0.14 | 0.09 | 0.2 | 0.06 | 0.01 | 0.04 |
| CE(14:0) | 0.43 | 0.52 | 1.34 | 0.79 | 0.38 | 0.49 | 0.86 | 0.66 | 0.4 | 0.59 | 0.6 | 0.56 | 2.83 | 5.43 | 0.33 | 0.53 |
| CE(15:0) | 0.3 | 0.46 | 1.21 | 0.27 | 0.2 | 0.43 | 0.29 | 0.44 | 0.61 | 0.79 | 0.34 | 0.41 | 2.81 | 5.98 | 0.35 | 0.52 |

[23]

| Protocol | 1 | | 2 | | 3 | | 4 | | 5 | | 6 | | 7 | | 8 | |
|---|---|---|---|---|---|---|---|---|---|---|---|---|---|---|---|---|
| **Metabolite** | **mean** | **SD** | **mean** | **SD** | **mean** | **SD** | **mean** | **SD** | **mean** | **SD** | **mean** | **SD** | **mean** | **SD** | **mean** | **SD** |
| CE(16:0) | 1.89 | 0.41 | 7.24 | 6.53 | 1.95 | 0.51 | 1.79 | 0.74 | 2.42 | 0.93 | 2.73 | 1.84 | 8.46 | 12.05 | 2.95 | 0.7 |
| CE(17:0) | 0.54 | 0.68 | 0.62 | 0.77 | 1.97 | 3.18 | 0.84 | 0.69 | 0.69 | 1.17 | 0.56 | 0.51 | 2.34 | 3.32 | 1.13 | 1.08 |
| CE(18:0) | 0.46 | 0.3 | 2.27 | 1.81 | 0.54 | 0.25 | 0.55 | 0.33 | 0.64 | 0.39 | 1.68 | 0.86 | 2.19 | 1.52 | 0.74 | 0.34 |
| CE(18:1) | 3.42 | 5.73 | 14.27 | 11.48 | 3.63 | 4.24 | 1.58 | 0.86 | 1.08 | 1.09 | 7.74 | 3.46 | 10.54 | 6.7 | 3.41 | 5.19 |
| CE(18:2) | 19.17 | 37.51 | 10.08 | 8.1 | 15.31 | 18.23 | 8.86 | 7.53 | 7.42 | 2.82 | 17.39 | 27.95 | 10.66 | 8.25 | 20.47 | 33.07 |
| CE(20:0) | 114.61 | 134.42 | 1.03 | 2.17 | 3.83 | 5.36 | 0.76 | 1.52 | 4.73 | 8.5 | 2.04 | 1.99 | 1.9 | 1.94 | 26.77 | 38.09 |
| CE(20:4) | 3.2 | 7.05 | 1.11 | 1.07 | 2.48 | 3.19 | 0.98 | 0.7 | 1.18 | 0.83 | 2.92 | 5.12 | 1.23 | 1.13 | 2.73 | 4.04 |
| p-Cresol-SO4 | 13.52 | 17.39 | 4 | 4.06 | 4.4 | 5.73 | 4.66 | 6.26 | 3.96 | 3.97 | 4.34 | 4.68 | 2.18 | 2.39 | 3.71 | 4.78 |
| DG(14:0_14:0) | 0.49 | 0.36 | 0.02 | 0.05 | 0.04 | 0.04 | 0.05 | 0.08 | 0.08 | 0.07 | 0.03 | 0.03 | 0.09 | 0.09 | 0.2 | 0.25 |
| DG(14:1/18:1) | 0.11 | 0.22 | 0.09 | 0.18 | 0 | 0 | 0.13 | 0.26 | 0.1 | 0.31 | 0.6 | 0.74 | 0.43 | 0.68 | 0.13 | 0.27 |
| DG(16:0_16:1) | 1.53 | 0.94 | 1.88 | 1.1 | 1.28 | 0.62 | 1.66 | 0.37 | 1.7 | 1.31 | 1.87 | 1.15 | 1.63 | 1.09 | 1.75 | 0.72 |
| DG(16:0_18:1) | 1.59 | 1.27 | 19.62 | 13.33 | 1.16 | 0.59 | 8.85 | 9.22 | 3.87 | 2.61 | 14.75 | 11.24 | 26.79 | 18.04 | 2.96 | 0.43 |
| DG(16:0_18:2) | 1.98 | 2.64 | 21.69 | 21.96 | 0.51 | 0.86 | 9.07 | 7.12 | 2.91 | 2.52 | 23.34 | 27.19 | 24.66 | 20.36 | 2.76 | 1.61 |
| DG(16:1_18:1) | 3.41 | 1.52 | 4.28 | 2.97 | 3.07 | 2.64 | 5.96 | 4.46 | 6.56 | 6.51 | 3.22 | 1.49 | 4.12 | 1.64 | 3.64 | 2.48 |
| DG(16:1_18:2) | 0 | 0 | 1.23 | 0.86 | 0.08 | 0.23 | 0.6 | 0.64 | 0 | 0 | 0.81 | 1.02 | 1.11 | 1.02 | 0.24 | 0.48 |
| DG(17:0_18:1) | 0.25 | 0.5 | 1.17 | 0.73 | 0.25 | 0.55 | 0.5 | 0.54 | 0.26 | 0.4 | 0.85 | 0.7 | 1.37 | 0.58 | 0.56 | 0.67 |
| DG(18:1_18:1) | 1.5 | 1.24 | 24.06 | 26.92 | 0.57 | 0.27 | 9.71 | 12.72 | 2.25 | 1.14 | 23.05 | 23.67 | 42.67 | 35.34 | 2.55 | 1.46 |
| DG(18:1_18:2) | 6.18 | 9.17 | 79.17 | 115.8 | 2.21 | 1.65 | 26.61 | 30.16 | 6.8 | 5.76 | 92.59 | 122.91 | 78.57 | 90.47 | 7.8 | 7.37 |
| DG(18:1_18:3) | 0.74 | 1.18 | 7.32 | 6.03 | 1.36 | 0.77 | 4.64 | 5.2 | 1.21 | 1.19 | 14.33 | 20.17 | 8.1 | 9.52 | 1.35 | 2.33 |
| DG(18:1_20:0) | 0.26 | 0.41 | 1.51 | 1.03 | 0.14 | 0.27 | 0.74 | 0.66 | 0.47 | 0.6 | 1.31 | 1.05 | 1.38 | 0.74 | 0.23 | 0.47 |
| DG(18:1_20:4) | 0.13 | 0.4 | 0.94 | 0.97 | 0.06 | 0.19 | 0.48 | 0.48 | 0.76 | 1.33 | 1.02 | 0.79 | 1.23 | 0.72 | 0.44 | 0.52 |
| DG(18:1_22:6) | 0.73 | 0.6 | 1.49 | 0.35 | 0.28 | 0.62 | 1.1 | 0.63 | 1.14 | 0.97 | 1.15 | 0.73 | 1.59 | 1.2 | 0.79 | 0.78 |
| DG(18:2_18:2) | 13.16 | 23.09 | 119.43 | 175.62 | 3.28 | 3.53 | 55.71 | 75.38 | 10.47 | 8.7 | 154.81 | 212.67 | 131.03 | 173.81 | 11.1 | 12.61 |
| DG(18:2_18:3) | 0.3 | 0.59 | 6.02 | 4.96 | 0.63 | 0.76 | 4.92 | 5.97 | 1.11 | 1.18 | 10.77 | 15.91 | 4.42 | 5.09 | 1.13 | 1.92 |
| DG(18:2_20:0) | 0.05 | 0.1 | 0.38 | 0.57 | 0.08 | 0.12 | 0.22 | 0.23 | 0.06 | 0.17 | 0.55 | 0.65 | 0.48 | 0.46 | 0.03 | 0.1 |
| DG(18:3_18:3) | 0.06 | 0.13 | 1.88 | 2.55 | 0.29 | 0.52 | 2.62 | 5.01 | 0.43 | 0.69 | 6 | 11.1 | 2.39 | 4.23 | 0.65 | 1.6 |
| DG(18:3_20:2) | 0.53 | 0.79 | 0.49 | 0.77 | 0.67 | 1.02 | 0.72 | 1.09 | 0.93 | 1.39 | 0.8 | 1.24 | 1.01 | 1.46 | 0.65 | 0.92 |
| Cer(d18:0/18:0(OH)) | 1.12 | 0.75 | 2.89 | 1.31 | 2.6 | 1.08 | 2.24 | 1.3 | 3.2 | 1.58 | 2.58 | 0.86 | 3.03 | 1.43 | 2.99 | 1.95 |
| Cer(d18:0/24:1) | 0.16 | 0.2 | 0.43 | 0.2 | 0.05 | 0.15 | 0.24 | 0.23 | 0.07 | 0.21 | 0.31 | 0.12 | 0.34 | 0.17 | 0.04 | 0.13 |
| Arachidonic acid | 0.24 | 0.13 | 12.88 | 11.95 | 15.96 | 8.89 | 13.25 | 8.82 | 13.75 | 10.36 | 19.01 | 14.18 | 16.98 | 10.77 | 14.47 | 12.24 |

| | | | | | | | | | [24] | | | | | | | |
|---|---|---|---|---|---|---|---|---|---|---|---|---|---|---|---|---|
| **Protocol** | **1** | | **2** | | **3** | | **4** | | **5** | | **6** | | **7** | | **8** | |
| **Metabolite** | **mean** | **SD** | **mean** | **SD** | **mean** | **SD** | **mean** | **SD** | **mean** | **SD** | **mean** | **SD** | **mean** | **SD** | **mean** | **SD** |
| DHA | 0.16 | 0.08 | 6.79 | 6.97 | 8.93 | 6.22 | 7.66 | 6.62 | 7.77 | 6.83 | 9.37 | 7.35 | 9.23 | 6.79 | 6.99 | 6.17 |
| EPA | 0.08 | 0.03 | 1.42 | 1.28 | 2.4 | 1.31 | 1.8 | 1.1 | 1.8 | 1.26 | 2.52 | 1.75 | 2.11 | 1.17 | 1.92 | 1.42 |
| FA(12:0) | 27.69 | 4.75 | 109.58 | 83.79 | 64.86 | 22.61 | 79.77 | 31.04 | 89.26 | 30.82 | 116.79 | 143.42 | 115.23 | 93.47 | 89.42 | 26.16 |
| FA(14:0) | 150 | 35.43 | 677.22 | 715.41 | 408.67 | 172.9 | 488.56 | 281.99 | 534.56 | 176.2 | 894 | 1497.15 | 714.11 | 729.24 | 479.11 | 101.71 |
| FA(18:1) | 175.11 | 222.68 | 2015.22 | 1712.97 | 1681.56 | 2000.64 | 1453.56 | 1368.24 | 1733.89 | 1940.16 | 5638.67 | 6978.52 | 5626.33 | 6366.23 | 1538.67 | 1543.05 |
| FA(18:2) | 336.02 | 512.13 | 1475.67 | 1538.92 | 2092.33 | 2582.13 | 1721.67 | 1841.39 | 1837.22 | 2103.27 | 2738.22 | 3243.14 | 2346.44 | 2623.84 | 1744.11 | 2083.99 |
| FA(20:1) | 4.09 | 1.36 | 65.59 | 31.68 | 15.72 | 8.3 | 27.89 | 12.7 | 33.09 | 22.18 | 174.41 | 200.16 | 166.14 | 180.77 | 24.99 | 7.72 |
| FA(20:2) | 2.99 | 0.7 | 19.15 | 9.35 | 13.57 | 3.3 | 15.82 | 7.31 | 20.07 | 5.23 | 19.32 | 7.78 | 21.67 | 10.12 | 16.24 | 4.44 |
| FA(20:3) | 1.57 | 1.11 | 17.96 | 18.88 | 13.84 | 9.43 | 17.09 | 14.45 | 15.14 | 12.78 | 18.82 | 13.32 | 19.44 | 15.08 | 17.69 | 18.29 |
| lysoPC a C16:0 | 0.83 | 0.28 | 5.04 | 3.07 | 6.6 | 3.18 | 5.72 | 2.71 | 6.09 | 2.13 | 4.84 | 3.15 | 8.14 | 3.15 | 5.52 | 2.96 |
| lysoPC a C17:0 | 0.06 | 0.07 | 0.23 | 0.16 | 0.21 | 0.13 | 0.26 | 0.13 | 0.25 | 0.2 | 0.18 | 0.13 | 0.27 | 0.12 | 0.26 | 0.16 |
| lysoPC a C18:0 | 0.32 | 0.14 | 2.77 | 1.6 | 2.14 | 0.89 | 2.8 | 1.51 | 2.91 | 1.21 | 1.87 | 1.22 | 3.6 | 1.61 | 2.7 | 1.53 |
| lysoPC a C18:1 | 0.11 | 0.11 | 1.04 | 0.89 | 2.31 | 2.25 | 1.55 | 1.54 | 1.43 | 0.93 | 1.01 | 0.48 | 9.61 | 18.41 | 1.41 | 0.97 |
| lysoPC a C18:2 | 0.15 | 0.13 | 0.78 | 0.61 | 2.66 | 2.93 | 1.48 | 1.12 | 1.69 | 1.94 | 1.08 | 0.77 | 1.96 | 1.97 | 1.17 | 0.8 |
| PC aa C24:0 | 0.07 | 0.11 | 0.1 | 0.16 | 0.06 | 0.04 | 0.04 | 0.04 | 0.07 | 0.05 | 0.11 | 0.2 | 0.04 | 0.06 | 0.11 | 0.06 |
| PC aa C30:0 | 0.18 | 0.02 | 0.67 | 0.46 | 0.22 | 0.02 | 0.33 | 0.09 | 0.39 | 0.06 | 0.29 | 0.14 | 0.55 | 0.19 | 0.42 | 0.1 |
| PC aa C32:0 | 0.14 | 0.08 | 1.31 | 0.91 | 0.14 | 0.04 | 0.44 | 0.36 | 0.4 | 0.19 | 0.44 | 0.26 | 1.35 | 0.68 | 0.41 | 0.27 |
| PC aa C32:1 | 0.04 | 0.04 | 0.29 | 0.21 | 0.06 | 0.07 | 0.24 | 0.13 | 0.15 | 0.08 | 0.13 | 0.1 | 0.51 | 0.17 | 0.16 | 0.13 |
| PC aa C32:2 | 0.02 | 0.06 | 0.11 | 0.08 | 0.01 | 0.02 | 0.05 | 0.06 | 0.05 | 0.05 | 0.04 | 0.04 | 0.16 | 0.05 | 0.1 | 0.1 |
| PC aa C32:3 | 0.01 | 0.02 | 0.05 | 0.04 | 0.01 | 0.02 | 0.01 | 0.02 | 0.01 | 0.02 | 0.01 | 0.02 | 0.06 | 0.04 | 0.02 | 0.03 |
| PC aa C34:1 | 1.43 | 0.46 | 3.36 | 2.63 | 1.5 | 0.82 | 2.1 | 0.99 | 1.68 | 0.29 | 1.35 | 1.06 | 6.91 | 5.6 | 2.48 | 1.46 |
| PC aa C34:2 | 0.99 | 1.91 | 1.83 | 0.85 | 0.41 | 0.49 | 1.35 | 0.64 | 0.76 | 0.21 | 0.67 | 0.76 | 3.54 | 2.56 | 0.59 | 0.62 |
| PC aa C34:3 | 0.01 | 0.03 | 0.24 | 0.16 | 0.06 | 0.05 | 0.19 | 0.15 | 0.16 | 0.24 | 0.14 | 0.15 | 0.37 | 0.25 | 0.11 | 0.09 |
| PC aa C34:4 | 0.01 | 0.01 | 0.03 | 0.04 | 0.01 | 0.02 | 0.02 | 0.03 | 0.03 | 0.04 | 0.02 | 0.02 | 0.05 | 0.04 | 0.02 | 0.03 |
| PC aa C36:1 | 0.15 | 0.12 | 0.76 | 0.58 | 0.21 | 0.27 | 0.33 | 0.23 | 0.19 | 0.09 | 0.25 | 0.22 | 1.41 | 0.54 | 0.23 | 0.2 |
| PC aa C36:2 | 2.34 | 0.94 | 2.61 | 1.88 | 2.24 | 0.73 | 1.91 | 0.32 | 2.67 | 0.47 | 1.95 | 1.39 | 19.45 | 46.85 | 2.86 | 0.8 |
| PC aa C36:3 | 0.08 | 0.11 | 1.43 | 1.9 | 0.22 | 0.17 | 1.05 | 0.82 | 0.52 | 0.49 | 0.49 | 0.71 | 3.13 | 2.84 | 0.57 | 0.77 |
| PC aa C36:4 | 0.12 | 0.13 | 0.91 | 0.74 | 0.41 | 0.42 | 0.95 | 0.78 | 0.49 | 0.52 | 0.5 | 0.97 | 3.28 | 5.67 | 0.24 | 0.29 |
| PC aa C36:5 | 0 | 0 | 0.24 | 0.32 | 0.07 | 0.07 | 0.24 | 0.4 | 0.24 | 0.45 | 0.22 | 0.42 | 0.41 | 0.56 | 0.04 | 0.04 |
| PC aa C36:6 | 0.01 | 0.02 | 0.07 | 0.1 | 0.05 | 0.03 | 0.11 | 0.14 | 0.1 | 0.21 | 0.07 | 0.09 | 0.13 | 0.16 | 0.05 | 0.03 |

[25]

| Protocol | 1 | | 2 | | 3 | | 4 | | 5 | | 6 | | 7 | | 8 | |
|---|---|---|---|---|---|---|---|---|---|---|---|---|---|---|---|---|
| **Metabolite** | **mean** | **SD** | **mean** | **SD** | **mean** | **SD** | **mean** | **SD** | **mean** | **SD** | **mean** | **SD** | **mean** | **SD** | **mean** | **SD** |
| PC aa C38:3 | 0.15 | 0.24 | 0.14 | 0.12 | 0.05 | 0.07 | 0.11 | 0.03 | 0.09 | 0.05 | 0.23 | 0.52 | 0.19 | 0.09 | 0.13 | 0.13 |
| PC aa C38:4 | 0.28 | 0.74 | 0.21 | 0.19 | 0.1 | 0.13 | 0.12 | 0.08 | 0.01 | 0.04 | 0.12 | 0.14 | 0.38 | 0.45 | 0.24 | 0.43 |
| PC aa C38:5 | 0.08 | 0.23 | 0.12 | 0.08 | 0.09 | 0.09 | 0.1 | 0.05 | 0.1 | 0.07 | 0.07 | 0.08 | 0.18 | 0.11 | 0.17 | 0.33 |
| PC aa C38:6 | 0.1 | 0.13 | 0.15 | 0.09 | 0.1 | 0.14 | 0.09 | 0.05 | 0.06 | 0.04 | 0.1 | 0.15 | 0.23 | 0.23 | 0.09 | 0.08 |
| PC aa C42:1 | 0.02 | 0.03 | 0.03 | 0.04 | 0.03 | 0.03 | 0.02 | 0.02 | 0.03 | 0.03 | 0.05 | 0.03 | 0.04 | 0.04 | 0.02 | 0.03 |
| PC aa C42:2 | 0.13 | 0.04 | 0.14 | 0.04 | 0.16 | 0.05 | 0.15 | 0.03 | 0.2 | 0.06 | 0.06 | 0.04 | 0.1 | 0.04 | 0.17 | 0.02 |
| PC ae C32:1 | 0.02 | 0.03 | 0.17 | 0.08 | 0.01 | 0.02 | 0.06 | 0.06 | 0.04 | 0.04 | 0.08 | 0.05 | 0.2 | 0.09 | 0.1 | 0.08 |
| PC ae C32:2 | 0.11 | 0.14 | 0.14 | 0.14 | 0.06 | 0.12 | 0.12 | 0.14 | 0.11 | 0.13 | 0.11 | 0.12 | 0.16 | 0.16 | 0.11 | 0.16 |
| PC ae C34:0 | 0.03 | 0.03 | 0.23 | 0.2 | 0.03 | 0.03 | 0.08 | 0.1 | 0.08 | 0.05 | 0.12 | 0.08 | 0.18 | 0.08 | 0.09 | 0.06 |
| PC ae C34:1 | 0.03 | 0.04 | 0.56 | 0.53 | 0.08 | 0.06 | 0.29 | 0.27 | 0.2 | 0.17 | 0.25 | 0.15 | 0.56 | 0.33 | 0.21 | 0.23 |
| PC ae C34:2 | 0.03 | 0.07 | 0.38 | 0.31 | 0.07 | 0.07 | 0.22 | 0.16 | 0.21 | 0.08 | 0.16 | 0.09 | 0.51 | 0.22 | 0.2 | 0.15 |
| PC ae C34:3 | 0.03 | 0.04 | 0.16 | 0.11 | 0.02 | 0.03 | 0.08 | 0.06 | 0.07 | 0.05 | 0.05 | 0.06 | 0.22 | 0.09 | 0.07 | 0.07 |
| PC ae C36:3 | 0 | 0.01 | 0.15 | 0.12 | 0.03 | 0.03 | 0.11 | 0.09 | 0.07 | 0.05 | 0.09 | 0.1 | 0.18 | 0.07 | 0.1 | 0.08 |
| PC ae C36:4 | 0.07 | 0.08 | 0.17 | 0.11 | 0.05 | 0.05 | 0.12 | 0.05 | 0.05 | 0.06 | 0.07 | 0.04 | 0.16 | 0.09 | 0.11 | 0.13 |
| PC ae C36:5 | 0 | 0.01 | 0.13 | 0.07 | 0.01 | 0.02 | 0.07 | 0.05 | 0.04 | 0.04 | 0.05 | 0.05 | 0.12 | 0.05 | 0.06 | 0.1 |
| PC ae C38:3 | 0.01 | 0.03 | 0.09 | 0.03 | 0.06 | 0.05 | 0.06 | 0.05 | 0.04 | 0.05 | 0.06 | 0.05 | 0.11 | 0.07 | 0.09 | 0.17 |
| PC ae C38:5 | 0.03 | 0.04 | 0.19 | 0.11 | 0.11 | 0.07 | 0.1 | 0.09 | 0.12 | 0.03 | 0.08 | 0.06 | 0.18 | 0.1 | 0.05 | 0.07 |
| PC ae C38:6 | 0.02 | 0.02 | 0.09 | 0.04 | 0.04 | 0.03 | 0.08 | 0.04 | 0.05 | 0.03 | 0.09 | 0.03 | 0.1 | 0.05 | 0.05 | 0.03 |
| Hex2Cer(d18:1/16:0) | 0.01 | 0.02 | 0.84 | 0.26 | 0 | 0 | 0.25 | 0.19 | 0.34 | 0.2 | 0.5 | 0.09 | 1.19 | 1 | 0.35 | 0.23 |
| Hex2Cer(d18:1/18:0) | 0.03 | 0.06 | 1.19 | 0.61 | 0.01 | 0.03 | 0.19 | 0.24 | 0.28 | 0.23 | 0.6 | 0.33 | 0.86 | 0.27 | 0.31 | 0.22 |
| Hex2Cer(d18:1/20:0) | 0 | 0.01 | 0.12 | 0.09 | 0 | 0 | 0 | 0 | 0.02 | 0.04 | 0.05 | 0.05 | 0.09 | 0.07 | 0 | 0 |
| Hex2Cer(d18:1/22:0) | 0.02 | 0.06 | 0.52 | 0.34 | 0 | 0 | 0.04 | 0.09 | 0.05 | 0.08 | 0.24 | 0.13 | 0.44 | 0.21 | 0.02 | 0.07 |
| Hex2Cer(d18:1/24:0) | 0 | 0 | 0.45 | 0.26 | 0.01 | 0.02 | 0.03 | 0.07 | 0.04 | 0.09 | 0.19 | 0.14 | 0.37 | 0.16 | 0.04 | 0.08 |
| Hex2Cer(d18:1/24:1) | 0 | 0 | 0.33 | 0.11 | 0 | 0 | 0.02 | 0.04 | 0.06 | 0.1 | 0.16 | 0.09 | 0.38 | 0.4 | 0.01 | 0.04 |
| Hex3Cer(d18:1/16:0) | 0 | 0 | 0.28 | 0.22 | 0 | 0 | 0 | 0 | 0.06 | 0.17 | 0.1 | 0.16 | 0.24 | 0.2 | 0.02 | 0.07 |
| Hex3Cer(d18:1/24:1) | 0.01 | 0.04 | 0.08 | 0.13 | 0 | 0 | 0 | 0 | 0.04 | 0.12 | 0.06 | 0.08 | 0.16 | 0.13 | 0 | 0 |
| Hex3Cer(d18:1/26:1) | 0 | 0 | 0 | 0 | 0.02 | 0.06 | 0 | 0 | 0 | 0 | 0.09 | 0.12 | 0.02 | 0.05 | 0 | 0 |
| Hex3Cer(d18:1/22:0) | 0 | 0 | 0.07 | 0.15 | 0 | 0 | 0.02 | 0.05 | 0 | 0 | 0.07 | 0.13 | 0 | 0 | 0.03 | 0.09 |
| HexCer(d16:1/22:0) | 0 | 0 | 0 | 0 | 0 | 0 | 0.05 | 0.08 | 0 | 0 | 0.07 | 0.12 | 0.03 | 0.07 | 0 | 0 |
| HexCer(d18:1/14:0) | 0 | 0 | 0.06 | 0.1 | 0 | 0 | 0.03 | 0.07 | 0.09 | 0.11 | 0.16 | 0.19 | 0.11 | 0.14 | 0.05 | 0.09 |

| Protocol | 1 | | 2 | | 3 | | 4 | | 5 | | 6 | | 7 | | 8 | |
|---|---|---|---|---|---|---|---|---|---|---|---|---|---|---|---|---|
| **Metabolite** | **mean** | **SD** | **mean** | **SD** | **mean** | **SD** | **mean** | **SD** | **mean** | **SD** | **mean** | **SD** | **mean** | **SD** | **mean** | **SD** |
| HexCer(d18:1/16:0) | 0 | 0 | 1.78 | 0.58 | 0.03 | 0.08 | 0.6 | 0.28 | 1 | 0.85 | 1.52 | 0.77 | 1.89 | 1.1 | 0.78 | 0.47 |
| HexCer(d18:1/18:0) | 0.02 | 0.05 | 0.66 | 0.68 | 0.01 | 0.03 | 0.11 | 0.16 | 0.2 | 0.33 | 0.41 | 0.18 | 0.47 | 0.43 | 0.07 | 0.11 |
| HexCer(d18:1/18:1) | 0.02 | 0.05 | 0.21 | 0.15 | 0.02 | 0.05 | 0.12 | 0.14 | 0.17 | 0.23 | 0.28 | 0.28 | 0.36 | 0.44 | 0.09 | 0.15 |
| HexCer(d18:1/20:0) | 0.02 | 0.07 | 0.38 | 0.4 | 0 | 0 | 0.04 | 0.07 | 0.13 | 0.38 | 0.2 | 0.22 | 0.3 | 0.3 | 0.07 | 0.1 |
| HexCer(d18:1/22:0) | 0.02 | 0.05 | 2.11 | 1.94 | 0.02 | 0.07 | 0.12 | 0.24 | 0.15 | 0.3 | 1.34 | 0.64 | 1.51 | 1.02 | 0.14 | 0.27 |
| HexCer(d18:1/23:0) | 0.11 | 0.22 | 1.08 | 0.8 | 0.04 | 0.11 | 0.12 | 0.26 | 0.54 | 0.64 | 1.03 | 0.32 | 0.78 | 0.56 | 0.31 | 0.42 |
| HexCer(d18:1/24:0) | 0.01 | 0.04 | 0.83 | 0.71 | 0.02 | 0.07 | 0.03 | 0.08 | 0.26 | 0.27 | 0.33 | 0.29 | 0.48 | 0.45 | 0.07 | 0.13 |
| HexCer(d18:1/24:1) | 0.18 | 0.27 | 4.88 | 1.92 | 0.21 | 0.32 | 0.4 | 0.51 | 1.32 | 0.93 | 2.59 | 0.64 | 3.29 | 1.5 | 0.86 | 0.6 |
| HexCer(d18:1/26:0) | 0 | 0 | 0.18 | 0.18 | 0 | 0 | 0.03 | 0.07 | 0.03 | 0.1 | 0.18 | 0.08 | 0.24 | 0.17 | 0.05 | 0.11 |
| HexCer(d18:2/16:0) | 0 | 0 | 0.19 | 0.21 | 0.04 | 0.08 | 0.07 | 0.11 | 0.08 | 0.15 | 0.15 | 0.16 | 0.27 | 0.18 | 0.18 | 0.19 |
| HexCer(d18:2/18:0) | 0.06 | 0.13 | 0.04 | 0.13 | 0.02 | 0.07 | 0 | 0 | 0 | 0 | 0.04 | 0.07 | 0.14 | 0.19 | 0 | 0 |
| HexCer(d18:2/22:0) | 0.17 | 0.26 | 0.06 | 0.18 | 0.06 | 0.17 | 0.11 | 0.23 | 0.17 | 0.37 | 0.16 | 0.26 | 0.17 | 0.26 | 0.09 | 0.28 |
| AbsAcid | 0.11 | 0.04 | 0.16 | 0.11 | 0.18 | 0.11 | 0.18 | 0.12 | 0.2 | 0.13 | 0.12 | 0.11 | 0.17 | 0.15 | 0.23 | 0.1 |
| DHEAS | 1.89 | 1.52 | 3.92 | 2.98 | 4.88 | 4.03 | 4.52 | 3.62 | 5.48 | 4.32 | 4.2 | 3.3 | 4.8 | 4.28 | 5.05 | 3.86 |
| 3-IAA | 67.94 | 69.73 | 22.5 | 20.16 | 25.63 | 17.31 | 26.86 | 23.64 | 29.19 | 19.01 | 19.94 | 14.23 | 21.04 | 15.46 | 31.84 | 24.37 |
| 3-IPA | 27.5 | 27.64 | 7.94 | 7.17 | 9.84 | 8.35 | 9.46 | 8.62 | 11.85 | 10.53 | 8.33 | 7.88 | 9.95 | 10 | 10.23 | 7.44 |
| Ind-SO4 | 0.66 | 0.9 | 0.11 | 0.08 | 0.34 | 0.41 | 0.24 | 0.24 | 0.28 | 0.3 | 0.16 | 0.18 | 0.29 | 0.28 | 0.36 | 0.43 |
| Indole | 1280.44 | 1205.28 | 575.56 | 407.54 | 542.67 | 288.89 | 601.67 | 441.38 | 587.22 | 272.54 | 367.56 | 222.79 | 238.33 | 105.25 | 639.11 | 307.32 |
| Hypoxanthine | 243.56 | 39.59 | 96.2 | 45.2 | 167.8 | 119.51 | 129.51 | 92.08 | 125.54 | 61.11 | 331.22 | 204.7 | 101.73 | 79.9 | 142.54 | 112.94 |
| Xanthine | 328.34 | 245.49 | 134.56 | 111.19 | 253.79 | 287.25 | 231.08 | 296.86 | 178.09 | 135.76 | 306.68 | 286.09 | 166.02 | 136.65 | 244.32 | 269.49 |
| SM (OH) C16:1 | 0.01 | 0.02 | 0.06 | 0.07 | 0.02 | 0.04 | 0.02 | 0.03 | 0.03 | 0.04 | 0.03 | 0.04 | 0.06 | 0.05 | 0.01 | 0.01 |
| SM (OH) C22:2 | 0.04 | 0.08 | 0.03 | 0.04 | 0.01 | 0.02 | 0.01 | 0.02 | 0.02 | 0.02 | 0.02 | 0.03 | 0.02 | 0.02 | 0.06 | 0.08 |
| SM (OH) C24:1 | 0.01 | 0.02 | 0.05 | 0.04 | 0.01 | 0.02 | 0.02 | 0.02 | 0.01 | 0.02 | 0.03 | 0.03 | 0.03 | 0.02 | 0.02 | 0.03 |
| SM C16:0 | 0.31 | 0.65 | 1.9 | 1.04 | 0.11 | 0.16 | 0.49 | 0.31 | 0.69 | 0.24 | 0.9 | 0.63 | 1.56 | 0.75 | 0.91 | 0.59 |
| SM C18:0 | 0.05 | 0.09 | 0.49 | 0.33 | 0.02 | 0.03 | 0.09 | 0.07 | 0.1 | 0.09 | 0.19 | 0.16 | 0.31 | 0.16 | 0.12 | 0.1 |
| SM C18:1 | 0.01 | 0.02 | 0.02 | 0.04 | 0.01 | 0.02 | 0.01 | 0.02 | 0 | 0.01 | 0.03 | 0.04 | 0.03 | 0.03 | 0 | 0.01 |
| SM C22:3 | 0.02 | 0.06 | 0.02 | 0.03 | 0.06 | 0.13 | 0.13 | 0.11 | 0.11 | 0.2 | 0.04 | 0.07 | 0.07 | 0.08 | 0.14 | 0.2 |
| SM C24:1 | 0.07 | 0.13 | 0.2 | 0.16 | 0.06 | 0.06 | 0.06 | 0.06 | 0.12 | 0.07 | 0.11 | 0.12 | 0.18 | 0.1 | 0.14 | 0.1 |
| H1 | 3171.89 | 2071.93 | 881.56 | 227.8 | 1369.56 | 452.48 | 1023.56 | 222.23 | 1814.89 | 743.17 | 2703.89 | 801.69 | 666.44 | 245.48 | 1248.78 | 205.35 |

| Protocol | 1 | | 2 | | 3 | | 4 | | 5 | | 6 | | 7 | | 8 | |
|---|---|---|---|---|---|---|---|---|---|---|---|---|---|---|---|---|
| Metabolite | mean | SD | mean | SD | mean | SD | mean | SD | mean | SD | mean | SD | mean | SD | mean | SD |
| TG(14:0_34:1) | 0.05 | 0.15 | 2.29 | 4.23 | 0.03 | 0.09 | 0.16 | 0.25 | 0.11 | 0.22 | 1.21 | 1.26 | 5.45 | 9.07 | 0.4 | 0.79 |
| TG(14:0_34:2) | 0.12 | 0.24 | 0.88 | 1.15 | 0.03 | 0.1 | 0 | 0 | 0 | 0 | 0.58 | 0.45 | 1.91 | 2.08 | 0.18 | 0.31 |
| TG(14:0_36:1) | 0 | 0 | 0.34 | 0.77 | 0.04 | 0.13 | 0 | 0 | 0 | 0 | 0.04 | 0.13 | 1.06 | 1.72 | 0.05 | 0.14 |
| TG(14:0_36:2) | 0.15 | 0.23 | 1.64 | 2.64 | 0.05 | 0.1 | 0.04 | 0.13 | 0 | 0 | 0.98 | 0.6 | 4.09 | 6.04 | 0 | 0 |
| TG(14:0_36:3) | 0.23 | 0.29 | 0.61 | 0.94 | 0.04 | 0.11 | 0.05 | 0.16 | 0.03 | 0.1 | 0.75 | 0.86 | 2.06 | 2.22 | 0 | 0 |
| TG(14:0_36:4) | 0.17 | 0.44 | 0.41 | 0.47 | 0.02 | 0.05 | 0 | 0 | 0 | 0 | 0.52 | 0.94 | 1.06 | 1.18 | 0.02 | 0.07 |
| TG(16:0_28:1) | 0 | 0 | 1.39 | 2.34 | 0.04 | 0.11 | 0.19 | 0.39 | 0.05 | 0.14 | 0.93 | 0.98 | 2.79 | 4.81 | 0.11 | 0.32 |
| TG(16:0_28:2) | 0 | 0 | 0.46 | 0.78 | 0 | 0 | 0.2 | 0.4 | 0.17 | 0.5 | 0.26 | 0.4 | 0.7 | 0.97 | 0.14 | 0.29 |
| TG(16:0_30:2) | 0 | 0 | 1.52 | 3.45 | 0 | 0 | 0.27 | 0.55 | 0.24 | 0.71 | 0.89 | 1.18 | 3.48 | 6.89 | 0.1 | 0.31 |
| TG(16:0_32:0) | 4.35 | 0.7 | 3.87 | 0.57 | 3.89 | 0.61 | 4.09 | 0.85 | 5.37 | 1.3 | 7.08 | 6.84 | 12.58 | 19.17 | 5.35 | 0.69 |
| TG(16:0_32:1) | 0.49 | 0.47 | 3.37 | 4.66 | 0.31 | 0.39 | 0.52 | 0.45 | 0.43 | 0.57 | 1.73 | 1.3 | 5.96 | 9.68 | 1.01 | 1.12 |
| TG(16:0_32:2) | 0.23 | 0.24 | 2.1 | 1.76 | 0.21 | 0.26 | 0.21 | 0.21 | 0.03 | 0.1 | 0.9 | 0.75 | 2.63 | 2.34 | 0.32 | 0.57 |
| TG(16:0_33:1) | 0.14 | 0.42 | 0.7 | 0.72 | 0.17 | 0.4 | 0.53 | 0.72 | 0.61 | 0.99 | 0.38 | 0.47 | 1.05 | 0.51 | 0.91 | 1.32 |
| TG(16:0_33:2) | 0.08 | 0.17 | 0.41 | 0.34 | 0 | 0 | 0.15 | 0.22 | 0 | 0 | 0.24 | 0.21 | 0.89 | 0.62 | 0.13 | 0.39 |
| TG(16:0_34:0) | 2.9 | 0.46 | 3.1 | 1.33 | 2.69 | 0.56 | 2.95 | 0.44 | 3.93 | 0.9 | 2.62 | 1.15 | 8.3 | 8.01 | 3.54 | 0.52 |
| TG(16:0_34:1) | 2.16 | 1.82 | 28.75 | 32.27 | 1.24 | 0.54 | 1.75 | 1.47 | 1.94 | 3.02 | 16.26 | 9.09 | 59.71 | 79.25 | 2.69 | 1.84 |
| TG(16:0_34:2) | 2.49 | 3.52 | 24.73 | 22.75 | 0.73 | 0.73 | 0.74 | 0.62 | 0.98 | 0.67 | 17.87 | 16.46 | 41.75 | 43.36 | 1.83 | 1.4 |
| TG(16:0_34:3) | 0 | 0 | 2.64 | 2.11 | 0.04 | 0.12 | 0.06 | 0.18 | 0.1 | 0.21 | 4.02 | 5.54 | 3.4 | 3.44 | 0.18 | 0.38 |
| TG(16:0_35:1) | 0.07 | 0.13 | 0.61 | 0.22 | 0.08 | 0.17 | 0.11 | 0.17 | 0.04 | 0.11 | 0.39 | 0.37 | 0.88 | 0.83 | 0.3 | 0.42 |
| TG(16:0_35:2) | 0.02 | 0.07 | 0.46 | 0.3 | 0 | 0 | 0.06 | 0.12 | 0.06 | 0.19 | 0.24 | 0.43 | 0.89 | 0.93 | 0.09 | 0.21 |
| TG(16:0_35:3) | 0 | 0 | 0.18 | 0.29 | 0 | 0 | 0.02 | 0.07 | 0 | 0 | 0.36 | 0.36 | 0.28 | 0.35 | 0 | 0 |
| TG(16:0_36:2) | 4.61 | 5.09 | 63.53 | 77.63 | 1.7 | 0.8 | 1.76 | 1.08 | 2.38 | 1.69 | 43.77 | 42.88 | 192.43 | 204.55 | 4.71 | 5.42 |
| TG(16:0_36:3) | 7.96 | 11.62 | 63.27 | 87.89 | 1.54 | 1.41 | 1.42 | 0.45 | 2.21 | 2.47 | 61.46 | 79.47 | 118.09 | 143.25 | 5.52 | 6.48 |
| TG(16:0_36:4) | 8.36 | 15.8 | 46.7 | 61.68 | 0.67 | 0.68 | 0.89 | 0.66 | 1.32 | 2.09 | 60.3 | 80.41 | 85.73 | 125.56 | 3.74 | 4.34 |
| TG(16:0_36:5) | 0 | 0 | 5.65 | 6.21 | 0 | 0 | 0.04 | 0.11 | 0.38 | 0.76 | 13.43 | 23.95 | 6.21 | 8.3 | 0.4 | 0.82 |
| TG(16:0_36:6) | 0.14 | 0.32 | 5.57 | 9.58 | 0.13 | 0.4 | 0.48 | 0.63 | 0.81 | 1.5 | 21.94 | 40.99 | 10.46 | 18.15 | 0.76 | 1.2 |
| TG(16:0_37:3) | 0 | 0 | 0.5 | 0.59 | 0 | 0 | 0 | 0 | 0 | 0 | 0.66 | 0.83 | 1.03 | 1.33 | 0 | 0 |
| TG(16:0_38:1) | 0 | 0 | 0.48 | 0.89 | 0 | 0 | 0 | 0 | 0.03 | 0.1 | 0.34 | 0.5 | 0.94 | 1.12 | 0.03 | 0.1 |
| TG(16:0_38:2) | 0 | 0 | 0.88 | 1.29 | 0 | 0 | 0.02 | 0.05 | 0 | 0 | 0.68 | 0.71 | 1.55 | 1.74 | 0.05 | 0.16 |
| TG(16:0_38:3) | 0.08 | 0.24 | 0.6 | 0.74 | 0.14 | 0.29 | 0.13 | 0.31 | 0.13 | 0.16 | 0.53 | 0.6 | 1.01 | 0.96 | 0.09 | 0.19 |

| | | | | | | | | | [28] | | | | | | | |
|---|---|---|---|---|---|---|---|---|---|---|---|---|---|---|---|---|
| **Protocol** | **1** | | **2** | | **3** | | **4** | | **5** | | **6** | | **7** | | **8** | |
| **Metabolite** | **mean** | **SD** | **mean** | **SD** | **mean** | **SD** | **mean** | **SD** | **mean** | **SD** | **mean** | **SD** | **mean** | **SD** | **mean** | **SD** |
| TG(16:1_32:1) | 0.16 | 0.34 | 2.27 | 2.7 | 0.3 | 0.35 | 0.19 | 0.28 | 0.19 | 0.41 | 0.88 | 1.46 | 1.77 | 1.69 | 1.37 | 2.45 |
| TG(16:1_32:2) | 0.12 | 0.24 | 4.15 | 5.76 | 0.04 | 0.11 | 0.4 | 0.46 | 0.09 | 0.21 | 1.62 | 2.65 | 2.7 | 3.25 | 0.49 | 1.16 |
| TG(16:1_34:1) | 0.31 | 0.53 | 2.86 | 2.87 | 0.45 | 0.4 | 0.38 | 0.37 | 0.09 | 0.26 | 1.34 | 1.33 | 3.9 | 2.89 | 0.8 | 0.98 |
| TG(16:1_34:2) | 0.21 | 0.42 | 5.47 | 6.27 | 0 | 0 | 0.12 | 0.27 | 0.07 | 0.22 | 2.28 | 2.84 | 4.09 | 4.02 | 0.45 | 0.82 |
| TG(16:1_34:3) | 0.03 | 0.08 | 0.63 | 0.72 | 0 | 0 | 0.03 | 0.08 | 0 | 0 | 0.26 | 0.41 | 0.56 | 0.58 | 0.16 | 0.37 |
| TG(16:1_36:1) | 0 | 0 | 1.46 | 1.09 | 0 | 0 | 0.06 | 0.11 | 0.08 | 0.25 | 0.55 | 0.46 | 2.33 | 1.69 | 0.16 | 0.34 |
| TG(16:1_36:2) | 0.2 | 0.59 | 3.56 | 2.91 | 0.09 | 0.17 | 0.09 | 0.14 | 0.16 | 0.31 | 1.95 | 1.16 | 11.16 | 11.59 | 0.35 | 0.59 |
| TG(16:1_36:3) | 0.44 | 0.56 | 2.13 | 1.54 | 0.1 | 0.3 | 0.05 | 0.11 | 0.06 | 0.13 | 1.95 | 1.69 | 3.94 | 3.36 | 0.26 | 0.43 |
| TG(16:1_36:4) | 0.26 | 0.64 | 1.41 | 1.27 | 0 | 0 | 0 | 0 | 0.02 | 0.05 | 1.59 | 1.6 | 2.18 | 2.56 | 0.07 | 0.22 |
| TG(16:1_38:3) | 0 | 0 | 0.29 | 0.5 | 0 | 0 | 0 | 0 | 0 | 0 | 0.23 | 0.42 | 0.6 | 0.63 | 0.03 | 0.09 |
| TG(16:1_38:4) | 0.04 | 0.12 | 0.38 | 0.75 | 0 | 0 | 0 | 0 | 0 | 0 | 0.55 | 0.83 | 0.7 | 0.98 | 0.05 | 0.14 |
| TG(16:1_38:5) | 0 | 0 | 0.22 | 0.38 | 0 | 0 | 0 | 0 | 0 | 0 | 0.35 | 0.53 | 0.43 | 0.69 | 0 | 0 |
| TG(17:0_36:3) | 0.02 | 0.07 | 0.43 | 0.68 | 0 | 0 | 0 | 0 | 0 | 0 | 0.45 | 0.59 | 0.94 | 1.09 | 0.11 | 0.22 |
| TG(17:0_36:4) | 0.08 | 0.12 | 0.28 | 0.47 | 0 | 0 | 0.02 | 0.06 | 0 | 0 | 0.47 | 0.63 | 0.66 | 0.82 | 0 | 0 |
| TG(17:1_34:2) | 0.03 | 0.09 | 0.27 | 0.56 | 0.06 | 0.13 | 0 | 0 | 0.08 | 0.15 | 0 | 0 | 0.4 | 0.45 | 0.09 | 0.27 |
| TG(17:1_36:3) | 0.02 | 0.06 | 0.27 | 0.53 | 0 | 0 | 0 | 0 | 0 | 0 | 0.12 | 0.26 | 0.7 | 0.8 | 0 | 0 |
| TG(17:1_36:4) | 0.07 | 0.22 | 0.25 | 0.41 | 0 | 0 | 0 | 0 | 0 | 0 | 0.31 | 0.48 | 0.45 | 0.65 | 0 | 0 |
| TG(17:2_36:2) | 0 | 0 | 0.11 | 0.23 | 0 | 0 | 0 | 0 | 0 | 0 | 0.21 | 0.32 | 0.63 | 0.68 | 0 | 0 |
| TG(17:1_36:3) | 0.02 | 0.05 | 0.2 | 0.4 | 0 | 0 | 0.02 | 0.06 | 0 | 0 | 0.33 | 0.49 | 0.52 | 0.49 | 0 | 0 |
| TG(17:2_36:4) | 0 | 0 | 0.4 | 0.52 | 0.03 | 0.08 | 0.1 | 0.15 | 0 | 0 | 0.43 | 0.54 | 0.57 | 0.68 | 0 | 0 |
| TG(18:0_30:1) | 0 | 0 | 0.81 | 1.9 | 0 | 0 | 0 | 0 | 0.04 | 0.11 | 0.48 | 0.64 | 1.99 | 4.17 | 0.05 | 0.14 |
| TG(18:0_32:1) | 0 | 0 | 1.08 | 1.15 | 0 | 0 | 0 | 0 | 0.09 | 0.28 | 0.52 | 0.43 | 1.35 | 1.24 | 0.14 | 0.29 |
| TG(18:0_32:2) | 1.39 | 1.66 | 1.82 | 1.91 | 1.14 | 1.64 | 1.37 | 1.82 | 1.58 | 2.29 | 1.33 | 1.8 | 2.36 | 1.71 | 1.71 | 1.71 |
| TG(18:0_34:2) | 1.04 | 1.83 | 9.73 | 12.97 | 0.04 | 0.13 | 0.13 | 0.2 | 0.29 | 0.37 | 8.4 | 11.19 | 15.73 | 19.73 | 0.44 | 0.67 |
| TG(18:0_34:3) | 0.04 | 0.12 | 0.8 | 0.91 | 0 | 0 | 0.06 | 0.12 | 0.04 | 0.13 | 1.47 | 2.94 | 1.08 | 1.15 | 0 | 0 |
| TG(18:0_36:1) | 0.22 | 0.38 | 6.01 | 8.3 | 0.05 | 0.16 | 0.03 | 0.09 | 0.2 | 0.24 | 4.2 | 5.42 | 13.42 | 14.33 | 0.12 | 0.37 |
| TG(18:0_36:2) | 1.76 | 2.65 | 24.72 | 36.2 | 0.36 | 0.35 | 0.39 | 0.41 | 0.65 | 0.75 | 20.12 | 26.58 | 68.2 | 68.19 | 1.65 | 2.68 |
| TG(18:0_36:3) | 3.02 | 4.98 | 28.9 | 48.41 | 0.36 | 0.3 | 0.3 | 0.34 | 1.07 | 1.23 | 33.69 | 49.31 | 47.6 | 64.23 | 2.21 | 3.86 |
| TG(18:0_36:4) | 3.55 | 6.52 | 21.42 | 34.77 | 0.16 | 0.31 | 0.49 | 0.62 | 1.05 | 1.51 | 33.58 | 51.04 | 38.43 | 57.28 | 1.84 | 2.69 |
| TG(18:0_36:5) | 0 | 0 | 1.89 | 2.48 | 0 | 0 | 0 | 0 | 0.16 | 0.38 | 4.81 | 10.72 | 1.97 | 3.23 | 0.26 | 0.63 |

[29]

| Protocol | 1 | | 2 | | 3 | | 4 | | 5 | | 6 | | 7 | | 8 | |
|---|---|---|---|---|---|---|---|---|---|---|---|---|---|---|---|---|
| **Metabolite** | **mean** | **SD** | **mean** | **SD** | **mean** | **SD** | **mean** | **SD** | **mean** | **SD** | **mean** | **SD** | **mean** | **SD** | **mean** | **SD** |
| TG(18:1_26:0) | 0.12 | 0.25 | 13 | 24.7 | 0.32 | 0.33 | 2.46 | 3.56 | 1.82 | 1.2 | 7.77 | 8.11 | 26.16 | 48.49 | 0.98 | 0.9 |
| TG(18:1_28:1) | 0.04 | 0.11 | 1.71 | 2.19 | 0.12 | 0.18 | 0.33 | 0.36 | 0.27 | 0.35 | 1.13 | 0.7 | 3.18 | 4.25 | 0.09 | 0.26 |
| TG(18:1_30:0) | 0.19 | 0.28 | 3.73 | 7.01 | 0.14 | 0.29 | 0.37 | 0.43 | 0.54 | 0.51 | 1.89 | 2.05 | 7.36 | 14.88 | 0.62 | 0.72 |
| TG(18:1_30:1) | 0.17 | 0.28 | 5.97 | 14.02 | 0.04 | 0.12 | 0.72 | 1.61 | 0.18 | 0.44 | 3.3 | 4.43 | 14.72 | 31.48 | 0.38 | 0.52 |
| TG(18:1_30:2) | 0.03 | 0.1 | 1.26 | 2.68 | 0 | 0 | 0.22 | 0.56 | 0.11 | 0.22 | 0.79 | 0.99 | 3.34 | 6.44 | 0 | 0 |
| TG(18:1_32:0) | 1.57 | 0.5 | 11.33 | 13.55 | 1.24 | 0.55 | 1.21 | 0.23 | 2 | 0.58 | 7.17 | 3.77 | 23.15 | 31.15 | 2.24 | 0.61 |
| TG(18:1_32:1) | 0.5 | 0.87 | 4.82 | 5.81 | 0.34 | 0.37 | 0.38 | 0.42 | 0.33 | 0.4 | 2.03 | 1.72 | 10.86 | 14.42 | 0.85 | 1.11 |
| TG(18:1_32:2) | 0.34 | 0.31 | 2.91 | 2.41 | 0.04 | 0.13 | 0.09 | 0.18 | 0.05 | 0.15 | 1.58 | 1.11 | 3.43 | 2.08 | 0.3 | 0.5 |
| TG(18:1_32:3) | 0 | 0 | 0.1 | 0.2 | 0.03 | 0.09 | 0 | 0 | 0.03 | 0.09 | 0.41 | 0.32 | 0.17 | 0.3 | 0 | 0 |
| TG(18:1_33:0) | 0.05 | 0.14 | 0.38 | 0.24 | 0.02 | 0.07 | 0.02 | 0.07 | 0 | 0 | 0.17 | 0.21 | 0.84 | 0.54 | 0.18 | 0.3 |
| TG(18:1_33:1) | 0 | 0 | 0.81 | 0.83 | 0.03 | 0.1 | 0.04 | 0.12 | 0.24 | 0.3 | 0.19 | 0.23 | 1.55 | 1.07 | 0.31 | 0.94 |
| TG(18:1_33:2) | 0 | 0 | 0.42 | 0.58 | 0 | 0 | 0.03 | 0.1 | 0 | 0 | 0.43 | 0.41 | 0.78 | 0.52 | 0.14 | 0.3 |
| TG(18:1_34:1) | 6.29 | 5.98 | 98.63 | 119.7 | 2.39 | 1.19 | 2.95 | 2.52 | 4.37 | 4.93 | 60.99 | 54.4 | 319.18 | 358.1 | 7.65 | 8.1 |
| TG(18:1_34:2) | 7.48 | 9.73 | 61.19 | 78.33 | 1.33 | 0.99 | 1.41 | 0.81 | 1.76 | 1.13 | 53.94 | 66.85 | 122.16 | 127.99 | 5.03 | 5.99 |
| TG(18:1_34:3) | 0.85 | 0.87 | 6.13 | 5.6 | 0.11 | 0.24 | 0.39 | 0.42 | 0.47 | 0.74 | 12.2 | 18.75 | 10.92 | 12.56 | 0.84 | 0.49 |
| TG(18:1_34:4) | 0.08 | 0.16 | 0.35 | 0.44 | 0.04 | 0.12 | 0.03 | 0.09 | 0.12 | 0.23 | 0.71 | 0.85 | 0.76 | 0.64 | 0.03 | 0.1 |
| TG(18:1_35:2) | 0 | 0 | 1.5 | 1.5 | 0 | 0 | 0 | 0 | 0 | 0 | 0.87 | 0.6 | 3.97 | 5.03 | 0.11 | 0.22 |
| TG(18:1_35:3) | 0 | 0 | 0.44 | 0.59 | 0 | 0 | 0 | 0 | 0.01 | 0.04 | 0.37 | 0.49 | 0.8 | 0.79 | 0.07 | 0.14 |
| TG(18:1_36:0) | 0.4 | 0.42 | 5.91 | 7.99 | 0.45 | 0.2 | 0.29 | 0.28 | 0.56 | 0.46 | 4.17 | 4.75 | 16.32 | 16.6 | 0.74 | 0.39 |
| TG(18:1_36:1) | 3.6 | 3.62 | 64.64 | 81.39 | 0.82 | 0.5 | 1.32 | 0.58 | 3.03 | 3.33 | 43.71 | 48.12 | 252.6 | 279.32 | 4.54 | 6.74 |
| TG(18:1_36:2) | 16.04 | 17.17 | 304.54 | 398.47 | 3.36 | 0.94 | 8.2 | 9.13 | 14.26 | 22.06 | 203.51 | 199.85 | 1408.61 | 1554.68 | 19.72 | 27.72 |
| TG(18:1_36:3) | 21.05 | 35.89 | 218.5 | 319.36 | 1.94 | 0.77 | 2.84 | 1.46 | 6.99 | 8.85 | 224.62 | 314.38 | 454.12 | 469 | 14.08 | 23.55 |
| TG(18:1_36:4) | 22.9 | 42.71 | 125.47 | 199.17 | 1.03 | 0.7 | 2.52 | 2.59 | 5.27 | 7.21 | 177.95 | 262.06 | 236.54 | 344.2 | 9.1 | 13.42 |
| TG(18:1_36:5) | 1.15 | 2.15 | 10.05 | 12.34 | 0.19 | 0.3 | 0.32 | 0.5 | 1.04 | 1.84 | 23.26 | 38.29 | 16.84 | 25.71 | 1.22 | 1.51 |
| TG(18:1_36:6) | 0.19 | 0.39 | 5.28 | 9.81 | 0.09 | 0.17 | 0.22 | 0.66 | 1.56 | 3.05 | 25.91 | 51.9 | 11.31 | 19.91 | 1.06 | 2.15 |
| TG(18:1_38:7) | 0 | 0 | 0.18 | 0.35 | 0 | 0 | 0 | 0 | 0 | 0 | 0.18 | 0.25 | 0.43 | 0.52 | 0 | 0 |
| TG(18:2_28:0) | 0 | 0 | 2.22 | 4.63 | 0.05 | 0.1 | 0.21 | 0.63 | 0 | 0 | 1.06 | 1.51 | 4.28 | 8.75 | 0.12 | 0.35 |
| TG(18:2_30:0) | 0.04 | 0.11 | 0.91 | 1.78 | 0.14 | 0.23 | 0.1 | 0.19 | 0.11 | 0.24 | 0.57 | 0.55 | 2.06 | 3.57 | 0.14 | 0.3 |
| TG(18:2_30:1) | 0.03 | 0.08 | 1.25 | 2.93 | 0.11 | 0.16 | 0.31 | 0.65 | 0 | 0 | 0.82 | 1 | 3.37 | 6.86 | 0.17 | 0.34 |
| TG(18:2_32:0) | 1.98 | 1.31 | 9.76 | 8.03 | 1.25 | 1.37 | 1.17 | 1.03 | 1.05 | 1.42 | 7.33 | 5.6 | 16.32 | 17.03 | 1.7 | 1.12 |

| Protocol | 1 | | 2 | | 3 | | 4 | | 5 | | 6 | | 7 | | 8 | |
|---|---|---|---|---|---|---|---|---|---|---|---|---|---|---|---|---|
| **Metabolite** | **mean** | **SD** | **mean** | **SD** | **mean** | **SD** | **mean** | **SD** | **mean** | **SD** | **mean** | **SD** | **mean** | **SD** | **mean** | **SD** |
| TG(18:2_32:1) | 0.08 | 0.17 | 1.47 | 1.09 | 0.03 | 0.08 | 0.08 | 0.16 | 0.14 | 0.29 | 1.14 | 0.74 | 3.06 | 2.69 | 0.45 | 0.66 |
| TG(18:2_32:2) | 0.3 | 0.89 | 1.26 | 1.02 | 0 | 0 | 0.07 | 0.14 | 0 | 0 | 1.11 | 1.46 | 2.28 | 2.1 | 0.09 | 0.26 |
| TG(18:2_33:1) | 0 | 0 | 0.2 | 0.24 | 0.03 | 0.1 | 0 | 0 | 0.03 | 0.09 | 0.27 | 0.31 | 0.73 | 0.62 | 0.16 | 0.27 |
| TG(18:2_33:2) | 0 | 0 | 0.5 | 0.53 | 0 | 0 | 0 | 0 | 0.03 | 0.07 | 0.55 | 0.6 | 0.68 | 0.64 | 0.02 | 0.07 |
| TG(18:2_34:0) | 1.43 | 2.28 | 11.03 | 14.77 | 0.15 | 0.23 | 0.19 | 0.25 | 0.36 | 0.36 | 10.21 | 12.99 | 18.51 | 22.38 | 0.77 | 1.1 |
| TG(18:2_34:1) | 8.06 | 11.69 | 61 | 84.57 | 1.38 | 1.45 | 1.1 | 0.72 | 1.76 | 2.06 | 58.47 | 76.64 | 108.04 | 133.48 | 4.95 | 6.16 |
| TG(18:2_34:2) | 13.39 | 24.25 | 71.79 | 96.24 | 1.4 | 0.97 | 1.66 | 1.32 | 2.42 | 1.93 | 80.96 | 106.87 | 127.17 | 184.28 | 5.52 | 6 |
| TG(18:2_34:3) | 0.48 | 1.01 | 5.84 | 5.5 | 0.11 | 0.16 | 0.19 | 0.23 | 0.4 | 0.46 | 10.82 | 16.79 | 7.25 | 8.99 | 0.24 | 0.47 |
| TG(18:2_34:4) | 0 | 0 | 0.63 | 0.85 | 0.03 | 0.1 | 0 | 0 | 0.07 | 0.22 | 1.58 | 2.48 | 1.02 | 1.33 | 0.04 | 0.13 |
| TG(18:2_35:1) | 0.07 | 0.14 | 1.01 | 0.98 | 0 | 0 | 0 | 0 | 0 | 0 | 0.94 | 1 | 1.6 | 1.75 | 0.04 | 0.13 |
| TG(18:2_35:2) | 0 | 0 | 0.98 | 0.98 | 0.03 | 0.08 | 0 | 0 | 0 | 0 | 0.83 | 0.84 | 1.57 | 1.63 | 0.04 | 0.13 |
| TG(18:2_35:3) | 0 | 0 | 0.42 | 0.59 | 0 | 0 | 0 | 0 | 0 | 0 | 0.42 | 0.57 | 0.64 | 0.83 | 0.03 | 0.08 |
| TG(18:2_36:0) | 0.55 | 0.98 | 4.69 | 7.62 | 0.06 | 0.11 | 0 | 0 | 0.1 | 0.2 | 4.62 | 6.45 | 7.5 | 9.67 | 0.32 | 0.66 |
| TG(18:2_36:1) | 4.2 | 6.9 | 38.81 | 63 | 0.36 | 0.36 | 0.38 | 0.42 | 1.29 | 1.61 | 42.09 | 61.7 | 67.91 | 83.02 | 2.34 | 4.83 |
| TG(18:2_36:2) | 19.32 | 32.55 | 152.22 | 231.84 | 1.3 | 0.85 | 2.42 | 1.98 | 4.25 | 5.47 | 172.22 | 247.34 | 299.73 | 333.11 | 10.23 | 16.08 |
| TG(18:2_36:3) | 42.43 | 79.72 | 235.73 | 381.02 | 1.44 | 2.35 | 4.3 | 5.02 | 6.87 | 8.53 | 297.56 | 443.1 | 418.71 | 617.77 | 15.13 | 24.63 |
| TG(18:2_36:4) | 41.24 | 84.64 | 176.25 | 264.82 | 1.33 | 1.51 | 5.41 | 7.48 | 5.49 | 6.95 | 252.95 | 363.19 | 337.38 | 530.31 | 10.43 | 14.98 |
| TG(18:2_36:5) | 0.11 | 0.32 | 11.82 | 11.98 | 0.1 | 0.3 | 0.75 | 0.8 | 1.31 | 2.42 | 26.87 | 49.85 | 13.58 | 18.44 | 1.3 | 2.25 |
| TG(18:2_38:4) | 0 | 0 | 0.32 | 0.34 | 0 | 0 | 0 | 0 | 0 | 0 | 0.5 | 0.59 | 0.33 | 0.63 | 0 | 0 |
| TG(18:3_32:0) | 0.11 | 0.17 | 0.87 | 0.83 | 0.03 | 0.08 | 0.04 | 0.09 | 0 | 0 | 1.39 | 2.22 | 0.8 | 1.1 | 0.19 | 0.3 |
| TG(18:3_34:0) | 0.16 | 0.26 | 0.65 | 1.14 | 0 | 0 | 0 | 0 | 0 | 0 | 2.27 | 4.44 | 1.2 | 1.85 | 0.05 | 0.14 |
| TG(18:3_34:1) | 0.36 | 0.54 | 5.22 | 6.01 | 0.05 | 0.16 | 0 | 0 | 0.47 | 0.98 | 13.22 | 22.78 | 8.85 | 12.31 | 0.54 | 0.95 |
| TG(18:3_34:2) | 0 | 0 | 6.02 | 6.38 | 0.09 | 0.18 | 0.11 | 0.22 | 0.38 | 0.75 | 13.94 | 24.87 | 6.38 | 8.85 | 0.41 | 0.84 |
| TG(18:3_34:3) | 0.04 | 0.11 | 7.73 | 14.34 | 0 | 0 | 0.25 | 0.46 | 0.98 | 1.95 | 31.13 | 59.11 | 13.75 | 24.97 | 0.79 | 1.73 |
| TG(18:3_35:2) | 0.02 | 0.07 | 0.14 | 0.24 | 0 | 0 | 0 | 0 | 0 | 0 | 0.31 | 0.58 | 0.21 | 0.3 | 0 | 0 |
| TG(18:3_36:1) | 0.33 | 0.66 | 3.06 | 4.09 | 0 | 0 | 0 | 0 | 0.63 | 1.25 | 7.79 | 15 | 4.68 | 6.5 | 0.53 | 0.98 |
| TG(18:3_36:2) | 1.14 | 2.01 | 10.52 | 13.86 | 0.07 | 0.14 | 0.32 | 0.47 | 1.38 | 2.83 | 25 | 44.18 | 16.9 | 24.2 | 1.21 | 2.21 |
| TG(18:3_36:3) | 1.15 | 2.16 | 12.63 | 16.57 | 0.28 | 0.36 | 0.68 | 0.62 | 1.81 | 3.34 | 34.48 | 64.93 | 20.22 | 31.26 | 1.7 | 3.09 |
| TG(18:3_36:4) | 0.31 | 0.4 | 15.89 | 23.65 | 0.24 | 0.48 | 0.86 | 1.49 | 3.33 | 6.46 | 62.32 | 125.96 | 26.68 | 45.43 | 2.67 | 5.24 |
| TG(20:0_32:4) | 0 | 0 | 0.3 | 0.53 | 0 | 0 | 0 | 0 | 0 | 0 | 0.5 | 0.64 | 0.62 | 0.91 | 0 | 0 |

| Protocol | 1 | | 2 | | 3 | | 4 | | 5 | | 6 | | 7 | | 8 | |
|---|---|---|---|---|---|---|---|---|---|---|---|---|---|---|---|---|
| **Metabolite** | **mean** | **SD** | **mean** | **SD** | **mean** | **SD** | **mean** | **SD** | **mean** | **SD** | **mean** | **SD** | **mean** | **SD** | **mean** | **SD** |
| TG(20:0_34:1) | 0.25 | 0.31 | 0.84 | 0.91 | 0.1 | 0.22 | 0.12 | 0.24 | 0.45 | 0.58 | 0.58 | 0.59 | 1.56 | 1.31 | 0.12 | 0.36 |
| TG(20:1_34:1) | 0.11 | 0.34 | 0.57 | 0.57 | 0.03 | 0.1 | 0 | 0 | 0.04 | 0.13 | 0.39 | 0.41 | 0.93 | 0.82 | 0.03 | 0.08 |
| TG(20:1_34:2) | 0.15 | 0.29 | 0.33 | 0.66 | 0 | 0 | 0 | 0 | 0 | 0 | 0.46 | 0.56 | 0.94 | 0.99 | 0.07 | 0.15 |
| TG(20:2_34:2) | 0 | 0 | 0.25 | 0.49 | 0 | 0 | 0 | 0 | 0 | 0 | 0.38 | 0.58 | 0.64 | 0.62 | 0 | 0 |
| TG(20:2_34:3) | 0 | 0 | 0.38 | 0.66 | 0 | 0 | 0 | 0 | 0 | 0 | 0.3 | 0.61 | 0.54 | 0.86 | 0 | 0 |
| TG(20:2_34:4) | 0 | 0 | 0.37 | 0.57 | 0 | 0 | 0 | 0 | 0 | 0 | 0.44 | 0.67 | 0.55 | 0.91 | 0 | 0 |
| TG(20:3_32:2) | 1.03 | 1.27 | 0.6 | 0.92 | 0.88 | 0.96 | 0.89 | 1.14 | 0.75 | 1.33 | 0.75 | 0.9 | 0.93 | 1.42 | 0.71 | 1.08 |
| TG(20:3_34:2) | 0 | 0 | 0.26 | 0.52 | 0 | 0 | 0 | 0 | 0 | 0 | 0.31 | 0.63 | 0.44 | 0.75 | 0 | 0 |
| TG(20:3_34:3) | 0 | 0 | 0.24 | 0.48 | 0 | 0 | 0 | 0 | 0 | 0 | 0.41 | 0.58 | 0.4 | 0.85 | 0 | 0 |
| TG(20:5_36:3) | 0 | 0 | 0.08 | 0.17 | 0.02 | 0.06 | 0 | 0 | 0 | 0 | 0.31 | 0.46 | 0.4 | 0.41 | 0.04 | 0.12 |
| TG(22:2_32:4) | 0.16 | 0.32 | 0.48 | 0.61 | 0.04 | 0.11 | 0.2 | 0.32 | 0.38 | 0.78 | 0.67 | 1.01 | 0.92 | 1.24 | 0.13 | 0.27 |
| Choline | 22.1 | 9.47 | 8.82 | 2.03 | 14.9 | 7.02 | 12.94 | 4.28 | 15.2 | 3.72 | 20.32 | 5.92 | 13.08 | 4.82 | 17.57 | 7.16 |

Concentrations below the LOD are marked in grey.

**Abbreviations**: 3-IAA, 3-Indoleacetic acid; 3-IPA, 3-Indolepropionic acid; 5-AVA, 5-Aminovaleric acid; AABA, alpha-Aminobutyric acid; Abs Acid, Abscisic acid; AconAcid, Aconitic acid; Ac-Orn, Acetylornithine; ADMA, Asymmetric dimethylarginine; Ala, Alanine; alpha-AAA, alpha Aminoadipic acid; Arg, Arginine; Asn, asparagine; Asp, aspartate; BABA, beta-Aminobutyric acid; betaAla, beta-Alanine; C0, Carnitine; C7-DC, Pimeloylcarnitine; C8, Octanoylcarnitine; C9, Nonaylcarnitine; C10, Decanoylcarnitine; C10:1, Decenoylcarnitine; C10:2, Decadienoylcarnitine; C12, Dodecanoylcarnitine; C16, Hexadecanoylcarnitine; C16-1OH, Hydroxyhexadecanoylcarnitine; C16:2, Hexadecadienoylcarnitine; C18, Octadecanoylcarnitine; CA, Cholic acid; CDCA, Chenodeoxycholic acid; CE, Cholesteryl ester; Cer, Ceramide; Cit, Citrulline; Cys, Cysteine; DCA, Deoxycholic acid; DG, Diglycerides; DHA, Docosahexaenoic acid, DHEAS, Dehydroepiandrosteron sulfate; DiCA(12:0), Dodecanedioic acid; DOPA, Dihydroxyphenylalanine; EPA, Eicosapentaenoic acid; FA, Fatty acid; Fa(12:0), Lauric acid; FA(14:0), Myristic acid; FA(18:1), Octadecenoic acid; FA(18:2), Octdecadienoic acid; FA(20:1), Eicosenoic acid; FA(20:2), Eicosadienoic acid; FA(20:3), Eicosatrienoic acid; H1, Hexoses (including glucose); GABA, gamma-Aminobutyric acid; GCA, Glycocholic acid; GDCA, Glycodeoxycholic acid; GLCA, Glycolithocholic acid; GLCAS, Glycolithocholic acid sulfate; Gln, Glutamine; Glu, Glutamate; Gly, Glycine; GUDCAS, Glycoursodeoxycholic aicd; HArg, Homoarginine; HCys, Homocysteine; HexCer, Hexosylceramide; Hex2Cer, Dihexosylceramide; Hex3Cer, Trihexosylceramide; His, Histidine; Ile, Isoleucine; Leu, Leucine; lysoPC, Lysophosphatidylcholine; Met, Methionine; Met-So, Methionine sulfoxide; OH-GlutAcid, 3-Hydroxyglutaric acid; Orn, Ornithine; PC, Phosphatidylcholine; p-cresol SO4, p-Cresol sulfate; PEA, Phenylethylamine; Phe, Phenylalanine; Pro, Proline; SDMA, Symmetric dimethylarginine; Ser, Serine; SM, Sphingomyelin; Suc, Succinic acid; t4-OH-Pro, trans-4-Hydroxyproline; TCA, Taurocholic acid; TCDCA, Taurochenodeoxycholic acid; TDCA, Taurodeoxycholic acid; TG, Triglyceride; Thr, Threonine; TLCA, Taurolithocholic acid; TMAO, Trimethylamine N-oxide; TMCA, Tauromurocholic acid; Trp, Tryptophan; Tyr, Tyrosine; Val, Valine.

**Supplementary Table 2**. Concentrations of metabolites sums and ratios (in pmol/mg stool) and their standard deviations that were measured in at least one of the tested protocols >LOD.

| Protocol | 1 | | 2 | | 3 | | 4 | | 5 | | 6 | | 7 | | 8 | |
|---|---|---|---|---|---|---|---|---|---|---|---|---|---|---|---|---|
| **Metabolite** | **mean** | **SD** | **mean** | **SD** | **mean** | **SD** | **mean** | **SD** | **mean** | **SD** | **mean** | **SD** | **mean** | **SD** | **mean** | **SD** |
| CACT Deficiency (NBS) | 183.51 | 87.92 | 28.92 | 25.66 | 59.53 | 52.05 | 56.98 | 57.46 | 63.32 | 46.72 | 45.09 | 37.68 | 47.48 | 46.58 | 59.24 | 47.67 |
| CPT-1 Deficiency (NBS) | 0.01 | 0.01 | 0.09 | 0.09 | 0.04 | 0.04 | 0.06 | 0.07 | 0.04 | 0.05 | 0.06 | 0.06 | 0.09 | 0.1 | 0.04 | 0.04 |
| Asn Synthesis | 0.04 | 0.03 | 0.06 | 0.07 | 0.01 | 0.01 | 0.03 | 0.01 | 0.01 | 0 | 0.11 | 0.03 | 0.02 | 0 | 0.01 | 0 |
| Cys Synthesis | 0.14 | 0.09 | 0.2 | 0.08 | 0.27 | 0.13 | 0.24 | 0.11 | 0.32 | 0.1 | 0.06 | 0.04 | 0.25 | 0.09 | 0.34 | 0.14 |
| DLD (NBS) | 1.88 | 0.51 | 5.14 | 2.67 | 5.08 | 2.55 | 5.39 | 2.81 | 5.04 | 2.88 | 0.92 | 0.39 | 3.84 | 2.15 | 5.37 | 3.01 |
| Fischer Ratio | 1.83 | 0.12 | 1.68 | 0.33 | 1.68 | 0.25 | 1.74 | 0.31 | 1.77 | 0.28 | 2.07 | 0.24 | 1.78 | 0.26 | 1.58 | 0.25 |
| Glutaminase Activity | 23.49 | 11.28 | 18.82 | 11.82 | 40.17 | 22.24 | 32.33 | 23.13 | 50.09 | 20.82 | 13.19 | 2.96 | 33.31 | 14.87 | 49.66 | 18.51 |
| Gly Synthesis | 1.6 | 0.38 | 2.46 | 0.78 | 2.56 | 0.81 | 2.47 | 0.58 | 2.66 | 0.81 | 1.34 | 0.23 | 2.89 | 0.92 | 2.79 | 0.92 |
| GSH Constituents | 5717.89 | 2724.31 | 604 | 313.75 | 4196.78 | 3579.62 | 2102.78 | 2181.33 | 4455.22 | 2380.5 | 3324.89 | 1830.53 | 2756.44 | 1755.98 | 5401.89 | 3332.66 |
| MTHFR Deficiency (NBS) | 1.04 | 0.11 | 1.21 | 0.51 | 1.24 | 0.51 | 1.18 | 0.54 | 1.23 | 0.45 | 0.96 | 0.13 | 0.9 | 0.24 | 1.09 | 0.38 |
| PKU (NBS) | 1.36 | 0.09 | 2.44 | 0.75 | 2.23 | 0.5 | 2.26 | 0.57 | 2.25 | 0.54 | 1.21 | 0.12 | 2.22 | 0.42 | 2.45 | 0.72 |
| Ratio of Non-Essential to Essential AAs | 3.5 | 1.28 | 2.61 | 0.97 | 5.26 | 3.52 | 4.66 | 3.76 | 5.32 | 2.93 | 2.01 | 0.33 | 4.8 | 2.93 | 5.37 | 2.93 |
| Sum of AAs | 11975.44 | 3543.55 | 1699.78 | 663.65 | 7043 | 4537.4 | 3771.22 | 2759.34 | 7763.33 | 2779.96 | 10584.13 | 5442.73 | 5253.33 | 2134.23 | 9262.33 | 4070.49 |
| Sum of Aromatic AAs | 486.22 | 113.59 | 107.63 | 31.56 | 183.96 | 74.8 | 147 | 62.73 | 181.56 | 37.78 | 899.22 | 585.52 | 193.11 | 73.84 | 225.33 | 80.73 |
| Sum of Branched-Chain AAs | 891.44 | 245.83 | 179.81 | 66.24 | 300.22 | 112.02 | 249 | 97.04 | 320.22 | 84.34 | 1937.44 | 1398.22 | 337 | 126.72 | 354.44 | 138.47 |
| Sum of Essential AAs | 2688.44 | 574.87 | 488 | 201.01 | 1101.11 | 319.52 | 681.56 | 282.3 | 1295.22 | 306.98 | 3538.63 | 1719.5 | 985.89 | 403.19 | 1480.89 | 335.55 |
| Sum of Non-Essential AAs | 9287 | 3353.55 | 1211.67 | 507.99 | 5941.89 | 4410.85 | 3089.89 | 2630.4 | 6468 | 2797.78 | 7657.22 | 3994.23 | 4267.33 | 2065.19 | 7781.56 | 3988.97 |
| Sum of Solely Glucogenic AAs | 9840.44 | 3391.08 | 1324.67 | 534.14 | 6166.11 | 4442.1 | 3252.22 | 2659.79 | 6709 | 2789.69 | 8448.78 | 4460.67 | 4465.22 | 2076.06 | 8037.22 | 4013.8 |
| Sum of Solely Ketogenic AAs | 1436.33 | 287.82 | 227.98 | 128.82 | 627.56 | 181.09 | 317.33 | 163.68 | 805.56 | 214.7 | 1761.13 | 734.21 | 527.78 | 258.49 | 924.44 | 205.08 |
| Sum of Sulfur-Containing AAs | 255.78 | 40.79 | 46.82 | 16.08 | 101.16 | 34.63 | 68.6 | 31.22 | 113.97 | 28.64 | 410.78 | 262.46 | 76.78 | 28.68 | 115.07 | 25.76 |
| Valinemia (NBS) | 1.69 | 0.27 | 2.76 | 0.52 | 2.73 | 0.54 | 2.9 | 0.78 | 2.8 | 0.26 | 1.5 | 0.21 | 2.69 | 0.47 | 2.71 | 0.48 |
| Cit Synthesis | 6.8 | 5.28 | 9.83 | 6.02 | 9.81 | 6.29 | 17.29 | 15.66 | 7.97 | 5.39 | 17.76 | 16.82 | 12.42 | 8.96 | 5.94 | 3.52 |
| OTC Deficiency (NBS) | 0.26 | 0.2 | 0.2 | 0.2 | 0.25 | 0.27 | 0.16 | 0.17 | 0.27 | 0.26 | 0.16 | 0.17 | 0.21 | 0.22 | 0.32 | 0.3 |
| Ratio of HArg to ADMA | 0.63 | 0.28 | 0.48 | 0.18 | 0.48 | 0.18 | 0.34 | 0.12 | 0.48 | 0.15 | 0.66 | 0.25 | 0.44 | 0.19 | 0.51 | 0.19 |
| Ratio of HArg to SDMA | 0.98 | 0.35 | 0.56 | 0.15 | 0.68 | 0.17 | 0.5 | 0.14 | 0.68 | 0.11 | 0.95 | 0.2 | 0.58 | 0.08 | 0.76 | 0.24 |

| Protocol | 1 | | 2 | | 3 | | 4 | | 5 | | 6 | | 7 | | 8 | |
|---|---|---|---|---|---|---|---|---|---|---|---|---|---|---|---|---|
| **Metabolite** | **mean** | **SD** | **mean** | **SD** | **mean** | **SD** | **mean** | **SD** | **mean** | **SD** | **mean** | **SD** | **mean** | **SD** | **mean** | **SD** |
| Sum of Betaine-Related Metabolites | 9.72 | 8.48 | 6.37 | 5.34 | 7.83 | 5.41 | 7.68 | 5.64 | 9.39 | 7.53 | 6.49 | 4.69 | 6.46 | 3.99 | 10.36 | 8.32 |
| Sum of Dimethylated Arg | 1.89 | 0.67 | 0.85 | 0.46 | 1.6 | 0.35 | 1.21 | 0.4 | 1.94 | 0.65 | 1.32 | 0.41 | 1.51 | 0.54 | 2.1 | 0.52 |
| Polyamine Synthesis | 2.38 | 1.16 | 4.48 | 2.2 | 2.07 | 1.41 | 3.09 | 2.12 | 1.69 | 0.84 | 1.52 | 0.73 | 2.7 | 1.9 | 1.47 | 0.9 |
| Putrescine Synthesis | 0.41 | 0.13 | 1.3 | 0.42 | 0.72 | 0.35 | 1.18 | 0.64 | 0.59 | 0.22 | 0.51 | 0.15 | 0.81 | 0.56 | 0.52 | 0.36 |
| Sum of Aminobutyric Acids | 47.52 | 24.34 | 23.13 | 14.36 | 40.97 | 10.04 | 32.64 | 10.2 | 47.16 | 23.63 | 47 | 9.62 | 52.88 | 11.71 | 64.08 | 34.87 |
| Sarcosine Synthesis from Choline | 1.21 | 1.13 | 1.1 | 0.83 | 1.41 | 0.99 | 1.2 | 0.86 | 1.52 | 1.12 | 0.76 | 0.5 | 1.57 | 1.27 | 1.55 | 1.38 |
| 3-Met-His Synthesis | 0.1 | 0.04 | 0.13 | 0.04 | 0.15 | 0.06 | 0.15 | 0.06 | 0.15 | 0.07 | 0.1 | 0.03 | 0.14 | 0.05 | 0.13 | 0.05 |
| AABA Synthesis | 0.13 | 0.08 | 0.3 | 0.12 | 0.25 | 0.12 | 0.33 | 0.17 | 0.24 | 0.11 | 0.12 | 0.07 | 0.42 | 0.23 | 0.29 | 0.11 |
| Asymmetrical Arg Methylation | 0.01 | 0 | 6.04 | 16.83 | 0.05 | 0.03 | 0.25 | 0.23 | 0.03 | 0.01 | 0 | 0 | 0.01 | 0.01 | 0.02 | 0.01 |
| BABA Synthesis | 0 | 0 | 0 | 0 | 0 | 0 | 0 | 0 | 0 | 0 | 0 | 0 | 0 | 0 | 0 | 0 |
| CPS Deficiency (NBS) | 2.68 | 0.38 | 3.81 | 1.66 | 5.41 | 1.97 | 4.19 | 1.98 | 6.51 | 2.34 | 1.69 | 0.58 | 3.12 | 1.01 | 4.91 | 1.81 |
| Cystine Synthesis | 0.03 | 0.02 | 0.01 | 0 | 0.01 | 0 | 0 | 0 | 0 | 0 | 0.05 | 0.03 | 0.01 | 0.01 | 0.01 | 0 |
| DOPA Synthesis | 0 | 0 | 0.01 | 0 | 0.01 | 0 | 0.01 | 0.01 | 0.01 | 0 | 0 | 0 | 0 | 0 | 0.01 | 0 |
| GABR | 0.22 | 0.05 | 0.09 | 0.12 | 0.18 | 0.2 | 0.08 | 0.1 | 0.16 | 0.11 | 0.74 | 0.38 | 0.73 | 0.67 | 0.5 | 0.52 |
| HArg Synthesis | 0 | 0 | 0.01 | 0.01 | 0 | 0 | 0 | 0 | 0 | 0 | 0 | 0 | 0 | 0 | 0 | 0 |
| HCys Synthesis | 0.08 | 0.03 | 0.36 | 0.14 | 0.24 | 0.11 | 0.29 | 0.14 | 0.28 | 0.07 | 0.06 | 0.04 | 0.25 | 0.11 | 0.31 | 0.15 |
| Met Oxidation | 0.05 | 0.01 | 0.12 | 0.07 | 0.1 | 0.03 | 0.1 | 0.02 | 0.08 | 0.02 | 0.08 | 0.04 | 0.21 | 0.07 | 0.08 | 0.02 |
| NO-Synthase activity | 3.87 | 0.93 | 19.67 | 15.77 | 12.47 | 10.45 | 53.04 | 50.49 | 7.84 | 5.67 | 1.67 | 1.21 | 2.4 | 1.94 | 4.11 | 3.96 |
| Orn Synthesis | 0.91 | 0.58 | 4.3 | 5.6 | 2.65 | 2.75 | 6.8 | 7.87 | 1.76 | 1.5 | 0.14 | 0.07 | 0.38 | 0.38 | 1.22 | 1.26 |
| Ratio of Pro to Cit | 0.73 | 0.29 | 1.33 | 0.39 | 0.93 | 0.28 | 1.28 | 0.31 | 0.78 | 0.37 | 0.63 | 0.32 | 1.16 | 0.27 | 1.05 | 0.25 |
| Sarcosine Synthesis from Gly | 0.05 | 0.04 | 0.11 | 0.07 | 0.08 | 0.05 | 0.13 | 0.09 | 0.08 | 0.05 | 0.04 | 0.03 | 0.1 | 0.06 | 0.08 | 0.06 |
| Sum of Asym. and Sym. Arg Methylation | 0.02 | 0.01 | 10.23 | 28.48 | 0.08 | 0.05 | 0.4 | 0.35 | 0.04 | 0.02 | 0 | 0 | 0.02 | 0.01 | 0.03 | 0.02 |
| Symmetrical Arg Methylation | 0.01 | 0 | 4.18 | 11.64 | 0.03 | 0.02 | 0.15 | 0.12 | 0.02 | 0.01 | 0 | 0 | 0.01 | 0 | 0.01 | 0.01 |
| Taurine Synthesis | 0.71 | 0.37 | 1.78 | 1.24 | 1.26 | 0.94 | 1.8 | 1.57 | 0.92 | 0.66 | 1.41 | 0.91 | 1.62 | 1.52 | 0.91 | 0.7 |
| GABA Synthesis | 0.01 | 0.01 | 0.03 | 0.03 | 0.01 | 0.01 | 0.03 | 0.03 | 0.01 | 0.01 | 0.01 | 0.01 | 0.01 | 0.01 | 0.01 | 0.01 |
| Histamine Synthesis | 0.05 | 0.04 | 0.19 | 0.14 | 0.13 | 0.12 | 0.17 | 0.15 | 0.13 | 0.1 | 0.07 | 0.05 | 0.11 | 0.11 | 0.11 | 0.1 |

[34]

| Protocol | 1 | | 2 | | 3 | | 4 | | 5 | | 6 | | 7 | | 8 | |
|---|---|---|---|---|---|---|---|---|---|---|---|---|---|---|---|---|
| **Metabolite** | **mean** | **SD** | **mean** | **SD** | **mean** | **SD** | **mean** | **SD** | **mean** | **SD** | **mean** | **SD** | **mean** | **SD** | **mean** | **SD** |
| PEA Synthesis | 0 | 0 | 0 | 0 | 0 | 0 | 0 | 0 | 0 | 0 | 0 | 0 | 0 | 0 | 0 | 0 |
| Serotonin Synthesis | 0.02 | 0.01 | 0.32 | 0.69 | 0.08 | 0.09 | 0.16 | 0.25 | 1.54 | 4.45 | 0.02 | 0.02 | 0.05 | 0.04 | 0.06 | 0.05 |
| p-Cresol-SO4 Synthesis | 0.05 | 0.05 | 0.06 | 0.06 | 0.04 | 0.04 | 0.05 | 0.05 | 0.04 | 0.04 | 0.01 | 0.01 | 0.02 | 0.01 | 0.03 | 0.03 |
| Indole Synthesis | 24.67 | 16.06 | 173.87 | 349.12 | 44.21 | 30.57 | 76.38 | 86.25 | 619.93 | 1749.07 | 7.48 | 3.15 | 18.74 | 11.32 | 40.23 | 23.65 |
| 7a-Dehydroxylation of CA | 7.47 | 7.49 | 10.89 | 8.73 | 9.39 | 7.64 | 8.13 | 6.21 | 11.62 | 11.69 | 7.11 | 4.68 | 7.89 | 6.11 | 10.52 | 9.67 |
| GDCA Synthesis from CA | 0.01 | 0.01 | 0.02 | 0.02 | 0.04 | 0.03 | 0.04 | 0.04 | 0.03 | 0.03 | 0.05 | 0.04 | 0.05 | 0.05 | 0.03 | 0.03 |
| GLCA Synthesis from CDCA | 0.03 | 0.02 | 0.01 | 0.02 | 0.01 | 0.01 | 0.01 | 0.01 | 0.01 | 0.01 | 0.01 | 0.01 | 0.01 | 0.03 | 0.01 | 0.01 |
| Gly Conjugation of CA | 0.06 | 0.07 | 0.06 | 0.05 | 0.19 | 0.25 | 0.35 | 0.8 | 0.16 | 0.24 | 0.11 | 0.12 | 0.15 | 0.17 | 0.53 | 0.92 |
| Gly Conjugation of CDCA | 0.32 | 0.32 | 0.13 | 0.13 | 0.24 | 0.28 | 0.21 | 0.28 | 0.15 | 0.15 | 0.21 | 0.31 | 0.6 | 1.18 | 0.37 | 0.66 |
| Gly Conjugation of DCA | 0 | 0 | 0 | 0.01 | 0.01 | 0.01 | 0 | 0.01 | 0 | 0 | 0.01 | 0.02 | 0.01 | 0.01 | 0 | 0 |
| Gly Conjugation of Primary BAs | 0.11 | 0.12 | 0.08 | 0.06 | 0.21 | 0.23 | 0.29 | 0.57 | 0.15 | 0.16 | 0.14 | 0.18 | 0.24 | 0.25 | 0.4 | 0.63 |
| Primary BA Conjugation | 0.18 | 0.16 | 0.22 | 0.14 | 0.36 | 0.29 | 0.52 | 0.92 | 0.3 | 0.24 | 0.17 | 0.18 | 0.42 | 0.34 | 0.69 | 0.88 |
| Ratio of 12a-OH BAs to Non-12a-O | 30.06 | 23.76 | 13.16 | 14.75 | 5.98 | 3.46 | 7.38 | 5.62 | 9.19 | 4.51 | 5.21 | 2.5 | 6.51 | 4.63 | 8.88 | 4.5 |
| Ratio of CDCA to CA | 0.21 | 0.12 | 0.95 | 0.61 | 1.31 | 0.62 | 1.26 | 0.83 | 1.03 | 0.61 | 1.59 | 1.2 | 1.17 | 0.79 | 1.08 | 0.73 |
| Ratio of Primary BAs to BAs | 0.28 | 0.17 | 0.27 | 0.18 | 0.34 | 0.18 | 0.33 | 0.12 | 0.31 | 0.17 | 0.35 | 0.18 | 0.34 | 0.15 | 0.31 | 0.15 |
| Ratio of Secondary BAs to BAs | 0.72 | 0.17 | 0.73 | 0.18 | 0.66 | 0.18 | 0.67 | 0.12 | 0.69 | 0.17 | 0.65 | 0.18 | 0.66 | 0.15 | 0.69 | 0.15 |
| Secondary BA Conjugation | 0.02 | 0.03 | 0.01 | 0.02 | 0.02 | 0.04 | 0.01 | 0.02 | 0.01 | 0 | 0.02 | 0.03 | 0.02 | 0.04 | 0.01 | 0.01 |
| Secondary BA Synthesis | 5.21 | 5.34 | 4.8 | 4.46 | 2.88 | 2.32 | 2.47 | 1.33 | 3.63 | 2.85 | 2.49 | 1.46 | 2.62 | 1.84 | 3.09 | 2.17 |
| Sum of 12a-OH BAs | 31.83 | 29.29 | 90.76 | 55.16 | 106.92 | 57.98 | 93.29 | 58.46 | 118.06 | 62.58 | 100 | 65.38 | 100.51 | 53.89 | 120.8 | 75.58 |
| Sum of BAs | 33.27 | 30.92 | 107.38 | 69.06 | 138.98 | 95.71 | 113.44 | 77.81 | 132.71 | 65.49 | 138.58 | 123.59 | 126.26 | 72.88 | 135.76 | 80.54 |
| Sum of Conjugated BAs | 0.51 | 0.21 | 6.49 | 10.23 | 12.07 | 18.69 | 8.08 | 11.67 | 5.58 | 2.26 | 8.56 | 16.24 | 12.38 | 15.88 | 7.58 | 3.62 |
| Sum of Conjugated Primary BAs | 0.41 | 0.22 | 5.47 | 8.71 | 10 | 14.86 | 6.74 | 9.26 | 4.82 | 1.8 | 6.73 | 12.8 | 10.2 | 12.15 | 6.77 | 3.41 |
| Sum of Conjugated Secondary BAs | 0.1 | 0.03 | 1.02 | 1.54 | 2.08 | 3.86 | 1.35 | 2.46 | 0.76 | 0.56 | 1.83 | 3.45 | 2.18 | 3.89 | 0.81 | 0.55 |
| Sum of Gly-Conjugated BAs | 0.22 | 0.07 | 2.18 | 3.61 | 4.48 | 5.15 | 2.9 | 3.49 | 2.2 | 1.15 | 5.31 | 8.4 | 5.52 | 6.47 | 3.31 | 2.52 |
| Sum of Non-12a-OH BAs | 1.44 | 1.86 | 16.67 | 21.89 | 32.44 | 49.71 | 20.2 | 24.66 | 14.59 | 8.47 | 38.53 | 64.07 | 25.84 | 26.64 | 14.97 | 7.8 |
| Sum of Primary BAs | 11.97 | 16.2 | 33.45 | 38.3 | 56.76 | 73.14 | 38.14 | 36.77 | 38.05 | 27.61 | 63.15 | 95.53 | 48.11 | 40.71 | 36.75 | 25.36 |

[35]

| Protocol | 1 | | 2 | | 3 | | 4 | | 5 | | 6 | | 7 | | 8 | |
|---|---|---|---|---|---|---|---|---|---|---|---|---|---|---|---|---|
| **Metabolite** | **mean** | **SD** | **mean** | **SD** | **mean** | **SD** | **mean** | **SD** | **mean** | **SD** | **mean** | **SD** | **mean** | **SD** | **mean** | **SD** |
| Sum of Secondary BAs | 21.29 | 16.4 | 73.92 | 43.26 | 82.53 | 44.44 | 75.32 | 48.38 | 94.77 | 55.36 | 75.24 | 40.14 | 78.33 | 41.65 | 99.13 | 61.29 |
| Sum of Taurine-Conjugated BAs | 0.28 | 0.25 | 4.31 | 6.64 | 7.59 | 13.72 | 5.18 | 8.34 | 3.38 | 1.82 | 3.24 | 7.88 | 6.87 | 9.73 | 4.28 | 2.26 |
| Sum of Unconjugated BAs | 32.75 | 30.72 | 100.9 | 63.16 | 127.23 | 80.79 | 105.17 | 69.79 | 127.02 | 64.21 | 129.79 | 108.61 | 114.06 | 63.28 | 128.17 | 81.96 |
| Sum of Unconjugated Primary BAs | 11.55 | 15.98 | 28 | 31.57 | 46.74 | 59.57 | 31.4 | 30.16 | 33.22 | 27.76 | 56.41 | 83.05 | 37.86 | 32.11 | 29.96 | 26.75 |
| Taurine Conjugation of CA | 0.06 | 0.04 | 0.1 | 0.1 | 0.15 | 0.12 | 0.27 | 0.46 | 0.18 | 0.16 | 0.02 | 0.02 | 0.19 | 0.14 | 0.38 | 0.39 |
| Taurine Conjugation of CDCA | 0.12 | 0.13 | 0.19 | 0.15 | 0.16 | 0.1 | 0.2 | 0.2 | 0.17 | 0.08 | 0.03 | 0.03 | 0.29 | 0.38 | 0.27 | 0.24 |
| Taurine Conjugation of DCA | 0 | 0 | 0.01 | 0.01 | 0.01 | 0.03 | 0.01 | 0.01 | 0 | 0 | 0.01 | 0.01 | 0.01 | 0.02 | 0 | 0 |
| Taurine Conjugation of Primary BAs | 0.07 | 0.05 | 0.13 | 0.11 | 0.15 | 0.1 | 0.23 | 0.35 | 0.15 | 0.11 | 0.02 | 0.02 | 0.19 | 0.12 | 0.29 | 0.27 |
| TDCA Synthesis from CA | 0.01 | 0.01 | 0.03 | 0.03 | 0.04 | 0.04 | 0.04 | 0.05 | 0.03 | 0.04 | 0.01 | 0.02 | 0.04 | 0.06 | 0.04 | 0.04 |
| TLCA Synthesis from CDCA | 0.02 | 0.01 | 0.02 | 0.02 | 0.02 | 0.02 | 0.01 | 0.01 | 0.02 | 0.02 | 0 | 0 | 0.03 | 0.03 | 0.02 | 0.02 |
| Spermidine Synthesis | 4.68 | 2.62 | 2.25 | 1.15 | 1.69 | 0.96 | 1.5 | 1.08 | 1.72 | 0.87 | 1.8 | 0.82 | 2.54 | 2.88 | 1.78 | 1.1 |
| Spermine Synthesis | 0.02 | 0 | 0.04 | 0.01 | 0.04 | 0.02 | 0.08 | 0.07 | 0.04 | 0.02 | 0.03 | 0.01 | 0.05 | 0.04 | 0.04 | 0.01 |
| Sum of Neurotransmitters | 6.24 | 3.92 | 6.49 | 4.67 | 7.4 | 5.38 | 7.06 | 5.35 | 8.82 | 6.22 | 6.18 | 5.13 | 6.27 | 5.64 | 8.47 | 5.09 |
| Sum of Polyamines | 199.56 | 32.17 | 58.1 | 28.34 | 68.34 | 29.84 | 44.97 | 28.35 | 80.78 | 23.32 | 62.32 | 14 | 37.64 | 14.11 | 73.2 | 20.18 |
| Ratio of DHA to arachidonic acid | 0.69 | 0.19 | 0.53 | 0.15 | 0.56 | 0.17 | 0.6 | 0.23 | 0.62 | 0.27 | 0.52 | 0.15 | 0.58 | 0.2 | 0.54 | 0.17 |
| Ratio of DHA to EPA | 2.21 | 1.54 | 4.27 | 1.14 | 3.71 | 1.42 | 3.91 | 1.49 | 4 | 1.41 | 3.82 | 1.44 | 4.26 | 1.71 | 3.49 | 1.4 |
| Ratio of EPA to arachidonic acid | 0.39 | 0.18 | 0.14 | 0.08 | 0.2 | 0.17 | 0.19 | 0.14 | 0.2 | 0.16 | 0.17 | 0.12 | 0.2 | 0.21 | 0.2 | 0.16 |
| Sum of Measured w-3 FAs | 0.25 | 0.08 | 8.21 | 8.25 | 11.32 | 7.38 | 9.47 | 7.7 | 9.57 | 8.06 | 11.9 | 9.06 | 11.34 | 7.86 | 8.92 | 7.55 |
| Sum of MUFAs | 179.16 | 223.78 | 2080.67 | 1740.54 | 1697.11 | 2008.3 | 1481.67 | 1379.06 | 1766.89 | 1961.04 | 5813.11 | 7176.29 | 5792.33 | 6536 | 1563.56 | 1547.5 |
| Sum of PUFAs | 341.11 | 511.67 | 1533.78 | 1514.79 | 2147 | 2569.65 | 1777.44 | 1820.37 | 1895.78 | 2080.4 | 2807.44 | 3223.52 | 2415.67 | 2596.57 | 1801.33 | 2059.17 |
| PLA2 Activity (2) | 8.17 | 4.79 | 27.39 | 21.48 | 142.43 | 174.12 | 32.93 | 26.21 | 61.59 | 54.08 | 87.08 | 98.55 | 20.84 | 13.44 | 36.36 | 16.72 |
| PLA2 Activity (4) | 4.4 | 5.15 | 72.94 | 45.12 | 177.34 | 113.4 | 116.2 | 90.77 | 146 | . | 210.27 | 206.24 | 71.65 | 54.96 | 55.85 | 31.17 |
| PLA2 Activity (6) | 6.05 | 3.6 | 85.9 | 43.56 | 259.58 | 200.62 | 169.8 | 72.74 | 387.53 | 355.39 | 269.38 | 202.93 | 98.99 | 52.4 | 115.7 | 71.54 |
| 3-IAA Synthesis | 0.05 | 0.01 | 0.04 | 0.01 | 0.05 | 0.01 | 0.04 | 0.01 | 0.05 | 0.01 | 0.05 | 0.01 | 0.08 | 0.03 | 0.05 | 0.01 |
| 3-IPA Synthesis | 0.02 | 0.01 | 0.01 | 0 | 0.02 | 0.01 | 0.01 | 0.01 | 0.02 | 0.01 | 0.02 | 0.01 | 0.03 | 0.02 | 0.01 | 0.01 |
| Sum of Purines | 571.89 | 262.17 | 230.78 | 152.22 | 421.67 | 401.15 | 360.56 | 377.11 | 303.67 | 187.07 | 638.11 | 487.11 | 267.78 | 210.21 | 386.96 | 369.77 |
| Xanthine Synthesis | 1.33 | 1.01 | 1.3 | 0.61 | 1.29 | 0.74 | 1.63 | 1.05 | 1.45 | 0.72 | 0.8 | 0.35 | 2.11 | 1.52 | 1.78 | 1.33 |

[36]

Concentrations below the LOD are marked in grey.

**Abbreviations**: 12a-OH BAs, 12-alpha hydroxylated bile acids, 3-IAA, 3-indoleacetic acid; 3-IPA, 3-indolepropionic acid; AA, amino acid; AABA, alpha-aminobutyric acid; ADMA, asymmetric dimethylarginine; Asn, asparagine; BA, bile acid; BABA, beta-aminobutyric acid; CA, cholic acid; CACT, carnitine-acylcarnitine translocase; CDCA, chenodeoxycholic acid; Cit, citrulline; CPS, carbamoyl phosphate synthetase; CPT1, carnitine palmitoyltransferase 1; Cys, cysteine; DCA, deoxycholic acid; DHA, docosahexaenoic acid; DLD, dihydrolipoamide dehydrogenase; DOPA, dihydroxyphenylalanine; EPA, eicosapentaenoic acid; FA, fatty acid; HipAcid, hippuric acid; GABA, gamma-aminobutyric acid; GABR, global arginine bioavailability ratio; Gly, glycine; GSH, glutathione; HArg, homoarginine; HCys, homocysteine; His, histidine; Met, methionine; MTHFR, methylene tetrahydrofolate reductase; MUFA, mono unsaturated fatty acid; NBS, newborn screening; NO, nitric oxide; Orn, ornithine; OTC, ornithine trancarbamylase; p-cresol SO4, p-cresol sulfate; PEA, phenylethylamine; PKU, phenylketonuria; PLA2, phospholipase 2; Pro, proline; PUFA, poly unsaturated fatty acid; SDMA, symmetric dimethylarginine; TDCA, taurodeoxycholic acid; TLCA, taurolithocholic acid; TMAO, trimethylamine N-oxide.
